# Effective coverage for maternal health: operationalizing effective coverage cascades for antenatal care and nutrition interventions for pregnant women in seven low- and middle-income countries

**DOI:** 10.1101/2024.06.29.24309704

**Authors:** Ashley Sheffel, Emily Carter, Rebecca Heidkamp, Aniqa Tasnim Hossain, Joanne Katz, Sunny Kim, Tsering Pema Lama, Tanya Marchant, Jamie Perin, Jennifer Requejo, Global Financing Facility, World Bank, Shelley Walton, Melinda K. Munos

**Author notes:** **Corresponding author:** Ashley Sheffel, Johns Hopkins University Bloomberg School of Public Health, Department of International Health 615 N Wolfe St., Baltimore, MD, USA 21205-2103.

## Abstract

**Background:** Efforts to improve maternal health have focused on measuring health and nutrition service coverage. However, high maternal mortality rates, despite improved service coverage, suggests that coverage indicators alone that do not account for quality can overestimate the health benefits of a service. Effective coverage (EC) cascades have been proposed as an approach to capture service quality within population-based coverage measures, but the proposed maternal health EC cascades have not been operationalized. This study aims to operationalize the effective coverage cascades for antenatal care (ANC) and maternal nutrition services using existing data from low- and middle-income countries (LMICs).

**Methods:** We used household surveys and health facility assessments from seven LMICs to estimate EC cascades for ANC and maternal nutrition services provided during ANC visits. We developed theoretical coverage cascades, defined health facility readiness and provision/experience of care scores and linked the facility-based scores to household survey data based on geographic domain and facility type. We then estimated the coverage cascade steps for each service by country.

**Findings:** Service contact coverage for at least one ANC visit (ANC1) was high, ranging from 80% in Bangladesh to 99% in Sierra Leone. However, there was a substantial drop in coverage from service contact to readiness-adjusted coverage, and a further drop to quality-adjusted coverage for all countries. For ANC1, from service contact to quality-adjusted coverage, there was an average net decline of 52 percentage points. For ANC1 maternal nutrition services, there was an average net decline of 48 percentage points from service contact to quality-adjusted coverage. This pattern persisted across cascades. Further exploration revealed that gaps in service readiness including lack of provider training, and gaps in provision/experience of care such as limited nutrition counseling were core contributors to the drops in coverage observed.

**Conclusions:** The cascade approach provided useful summary measures that identified major barriers to EC. However, detailed measures underlying the steps of the cascade are likely needed to support evidence-based decision-making with more actionable information. This analysis highlights the importance of understanding bottlenecks in achieving health outcomes and the inter-connectedness of service access and service quality to improve health in LMICs.

## BACKGROUND

Improving maternal health and survival in low-income and middle-income countries (LMICs) remains at the forefront of the global health and development agenda [1]. While considerable progress has been made over the last two decades, the majority of maternal morbidity and mortality is preventable, yet remains unacceptably high and disproportionately occurring in LMICs. In 2020, the maternal mortality ratio (MMR) for the world’s least developed countries was nearly 30 times than that of the subregion Europe and Northern America [2]. In addition, the persistently high prevalence of maternal malnutrition in LMICs has remained unacceptably high and has been exacerbated in recent years by the COVID-19 pandemic, increased conflict, and climate change resulting in further challenges to improving maternal health [3-6]. Key to improving maternal health is ensuring universal access to evidence-based interventions delivered with high quality [7-9].

Antenatal care (ANC) is an ideal platform on which to deliver and promote evidence-based, cost-effective interventions to improve maternal health within and beyond the pregnancy period. ANC is important to maintaining a healthy pregnancy [10-12], promotes safe delivery and postnatal attendance, and is positively associated with an increase in facility-based deliveries [13-15]. In addition, ANC provides women with an important contact with the formal health system, leading to opportunities to access and utilize evidence-based interventions which promote maternal health and survival [16,17] including the delivery of vital maternal nutrition interventions as part of a comprehensive ANC service [18].

Global and national efforts to monitor the implementation of evidence-based, cost-effective interventions to improve maternal health have focused on measuring health service contact coverage, defined as the proportion of the target population in need of a service that received the service [19]. However, evidence of persistently high maternal mortality levels despite considerable improvements in coverage suggests that service contact coverage alone (e.g., at least one ANC visit) without accounting for service quality [20,21] can overestimate the health benefits of a service.

The concept of effective coverage aims to move beyond coverage to generate a better estimate of the benefit of a health service. Effective coverage indicators estimate the proportion of a population in need of a service that received the service with sufficient quality to achieve a positive health outcome [22-25]. The global health community has reached a general consensus that effective coverage for maternal health interventions can be conceptualized using a coverage cascade framework which outlines six steps: 1) service contact coverage; 2) input-adjusted coverage; 3) intervention coverage; 4) quality-adjusted coverage (whereby quality refers to process quality or the provision and experience of care); 5) user adherence-adjusted coverage; and 6) outcome-adjusted coverage [22,23]. Effective coverage has been defined as outcome-adjusted coverage, the final step of the cascade. However, for some interventions, quality-adjusted coverage may be a more suitable proxy measurement of effective coverage, particularly for routine preventative or promotive health services such as ANC during which multiple interventions are delivered [22]. In addition, readiness- and quality-adjusted coverage can be influenced through health system strengthening whereas there are a multitude of factors beyond the health system that can impact health outcomes, making readiness- and quality-adjusted coverage measures particularly important for improving the health system.

Despite consensus within the global health community on the concept of effective coverage cascades, research on how to operationalize these cascades for various services and data sources is limited. A recent review of effective coverage of maternal, newborn, and child health (MNCH) interventions found no consistent approach to the adjustments made to contact coverage [26]. However, evidence is growing on best practices for defining and generating effective coverage cascades using data from various sources. Munos et al recently set forth a set of best practices for generating estimates for effective coverage cascades using both household survey (HHS) data and health facility assessment (HFA) data [27]. Exley et al have explored operationalizing the coverage cascade for facility-based childbirth interventions using HHS data and two sources of health facility data (HFA and routine health information data) [28] while Kim et al explored generating effective coverage estimates for maternal and newborn health services using HHS data [29]. The aim of this study was to operationalize effective coverage cascades for ANC and for maternal nutrition services delivered during ANC using extant nationally representative, publicly available data in LMICs. We aimed to provide guidance on defining the data source and content of each cascade step, linking data sources, and unique challenges in estimating the cascade for each service area.

## METHODS

### Overview

Using publicly available HHS data and HFA data from seven LMICs (specified below), we estimated effective coverage cascades for ANC and maternal nutrition interventions delivered through ANC visits. We developed theoretical coverage cascades for ANC and maternal nutrition, defined facility readiness and provision/experience of care scores for each service using facility survey data, linked those scores to HHS data based on household location and reported type of facility utilized, and estimated the steps of the coverage cascade for each service and country. We adhered to best practices for generating estimates for effective coverage cascades as detailed in Munos et al and further described below [27].

### Conceptualizing the effective coverage cascade

We utilized the coverage cascade framework proposed by Amouzou et al and adapted by the Effective Coverage Think Tank Group – a group of experts led by the World Health Organization (WHO) and the United Nations Children’s Fund (UNICEF) – to develop a theoretical care cascade for ANC and maternal nutrition (**Figure 1**) [22,23]. We defined effective coverage as the proportion of all pregnant women (the target population) who progressed through six steps: 1) attended a health facility for ANC (service contact coverage), 2) attended a health facility for ANC that had the appropriate inputs available (readiness-adjusted coverage), 3) received the appropriate interventions (intervention coverage), 4) attended a health facility for ANC where providers followed recommended standards or processes of care (quality-adjusted coverage), 5) adhered to selected interventions at home (user-adjusted coverage), and 6) had positive pregnancy outcomes (outcome-adjusted coverage).

**Figure 1:**
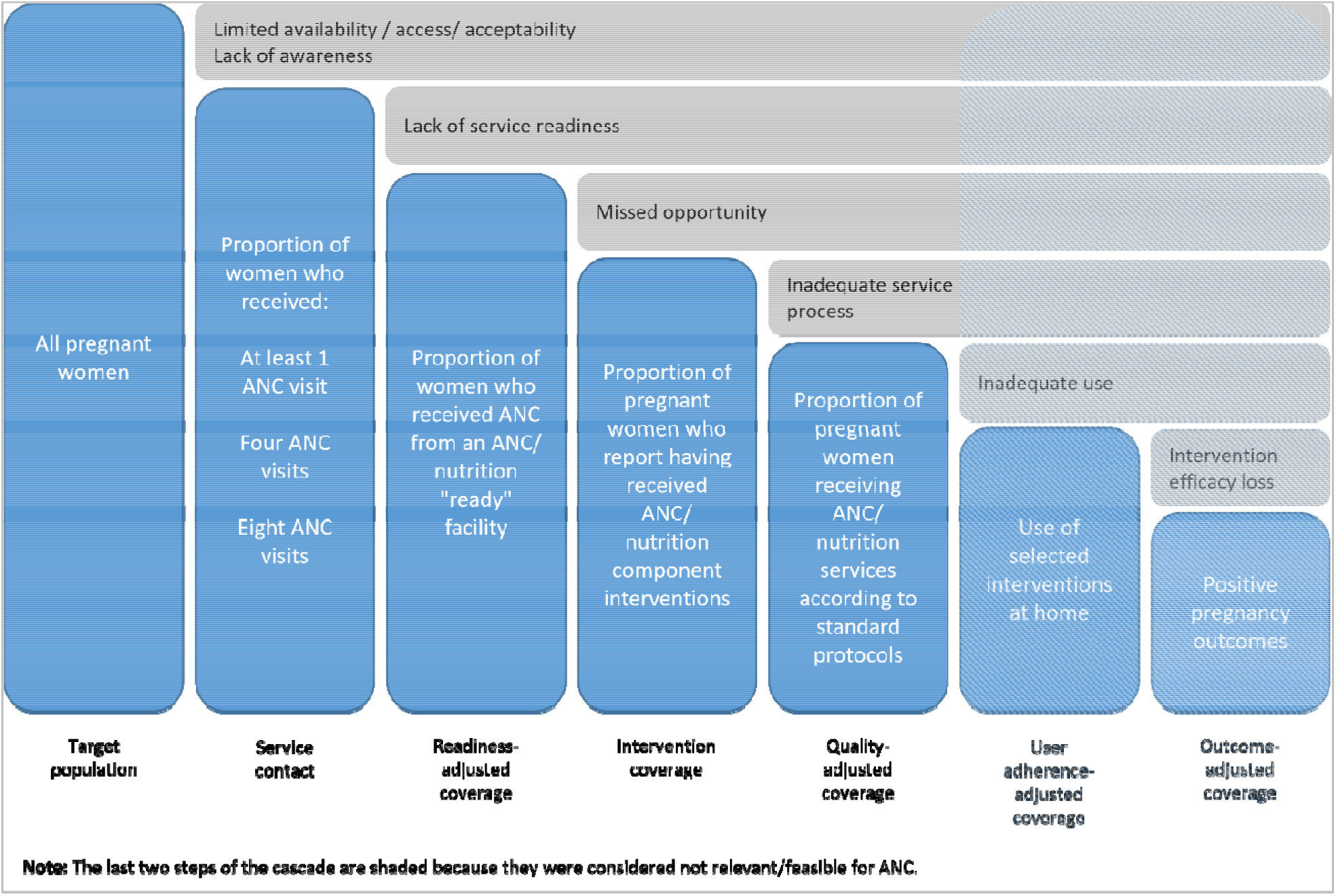
Theoretical effective coverage cascade for ANC and maternal nutrition.

**Table 1** provides operational definitions of each step of the coverage cascade for this analysis, using existing data from select countries. Further information on how to calculate each step of the cascade is provided subsequently in the methods section.

**Table 1:**
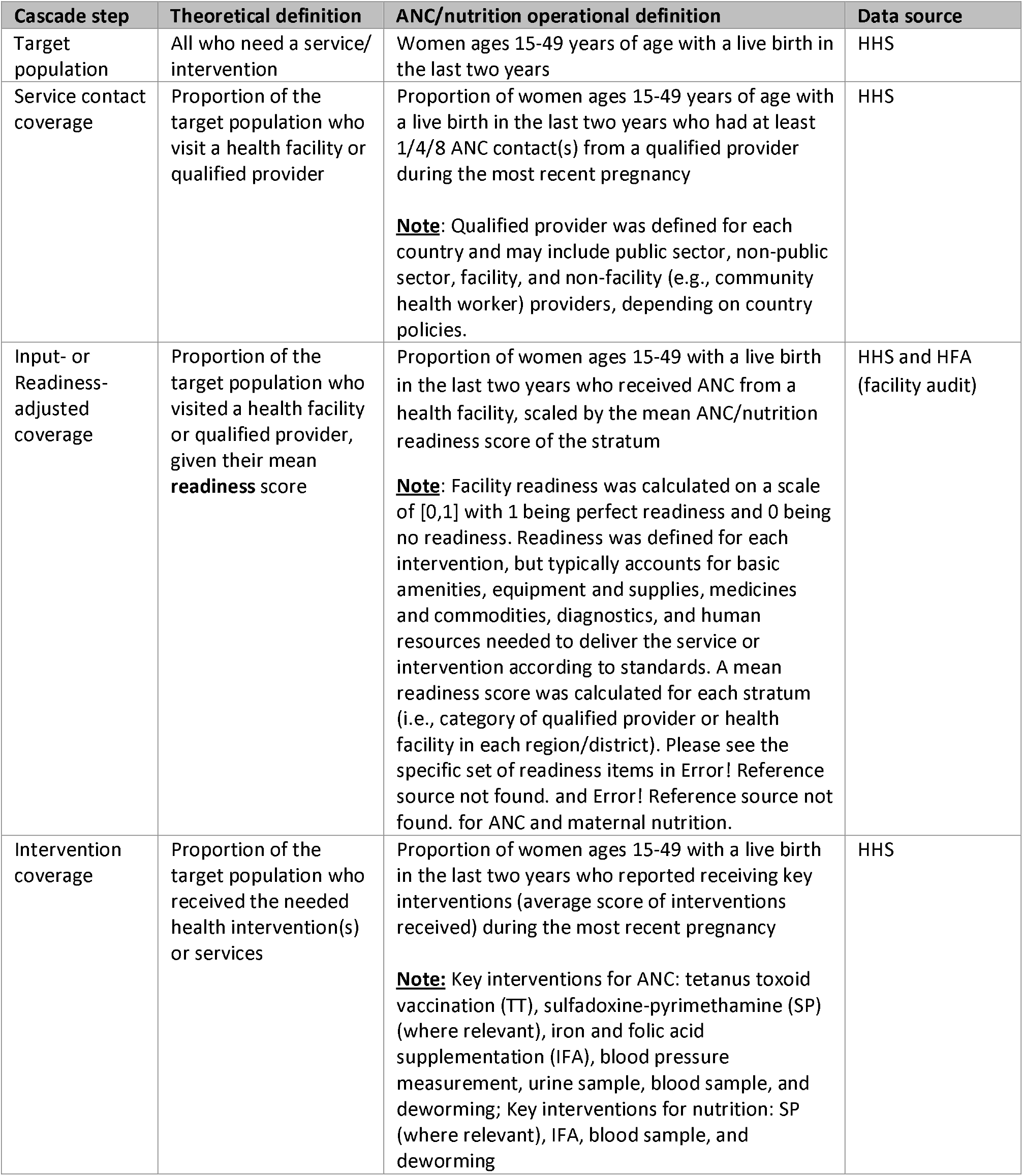
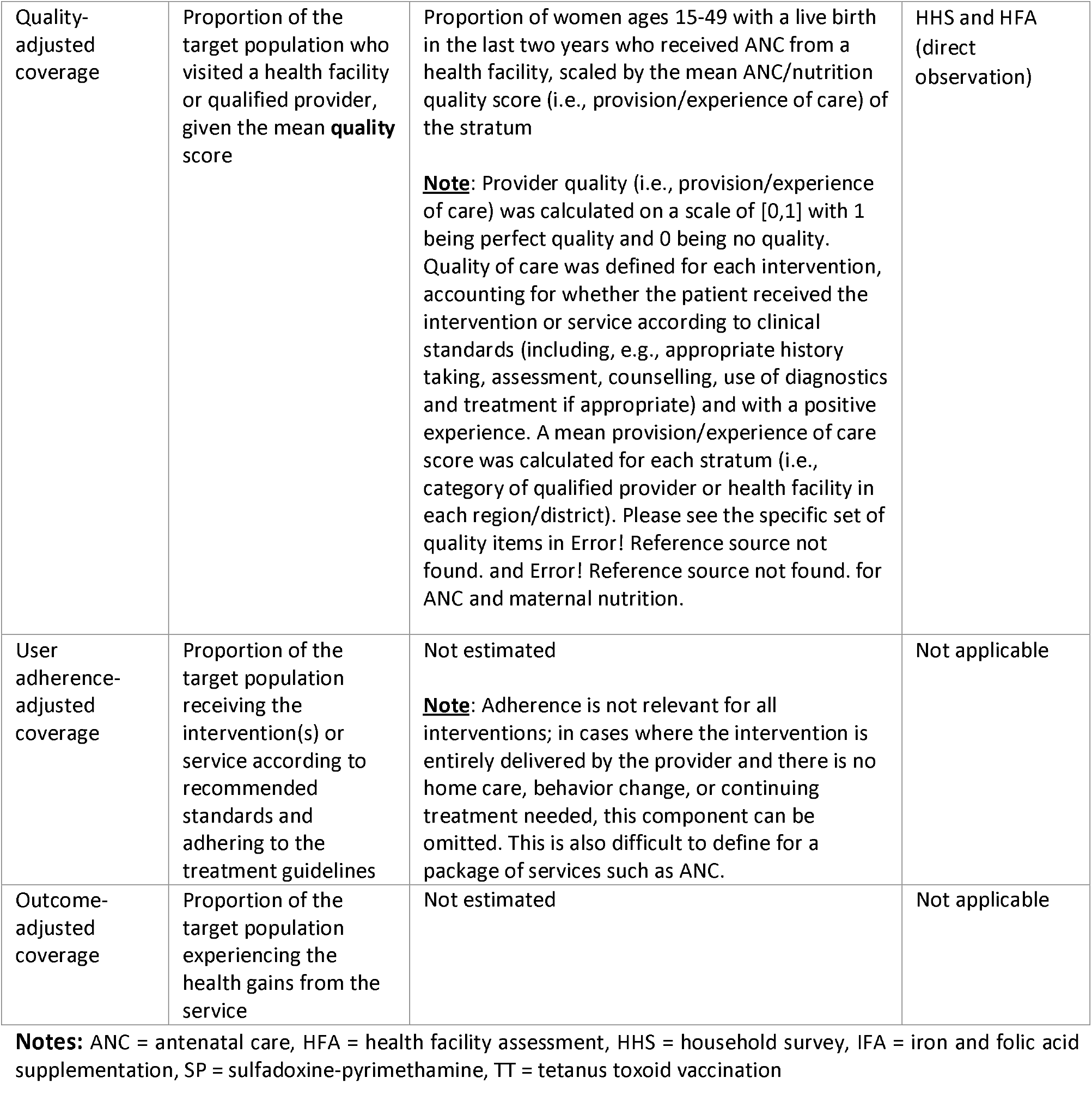
Operational definitions of the effective coverage cascade components.

| Cascade step | Theoretical definition | ANC/nutrition operational definition | Data source |
| --- | --- | --- | --- |
| Target population | All who need a service/ intervention | Women ages 15-49 years of age with a live birth in the last two years | HHS |
| Service contact coverage | Proportion of the target population who visit a health facility or qualified provider | Proportion of women ages 15-49 years of age with a live birth in the last two years who had at least 1/4/8 ANC contact(s) from a qualified provider during the most recent pregnancy<br>**Note:** Qualified provider was defined for each country and may include public sector, non-public sector, facility, and non-facility (e.g., community health worker) providers, depending on country policies. | HHS |
| Input- or Readiness-adjusted coverage | Proportion of the target population who visited a health facility or qualified provider, given their mean readiness score | Proportion of women ages 15-49 with a live birth in the last two years who received ANC from a health facility, scaled by the mean ANC/nutrition readiness score of the stratum<br>**Note:** Facility readiness was calculated on a scale of [0,1] with 1 being perfect readiness and 0 being no readiness. Readiness was defined for each intervention, but typically accounts for basic amenities, equipment and supplies, medicines and commodities, diagnostics, and human resources needed to deliver the service or intervention according to standards. A mean readiness score was calculated for each stratum (i.e., category of qualified provider or health facility in each region/district). Please see the specific set of readiness items in Error! Reference source not found. and Error! Reference source not found. for ANC and maternal nutrition. | HHS and HFA (facility audit) |
| Intervention coverage | Proportion of the target population who received the needed health intervention(s) or services | Proportion of women ages 15-49 with a live birth in the last two years who reported receiving key interventions (average score of interventions received) during the most recent pregnancy<br>**Note:** Key interventions for ANC: tetanus toxoid vaccination (TT), sulfadoxine-pyrimethamine (SP) (where relevant), iron and folic acid supplementation (IFA), blood pressure measurement, urine sample, blood sample, and deworming; Key interventions for nutrition: SP (where relevant), IFA, blood sample, and deworming | HHS |
| Quality-adjusted coverage | Proportion of the target population who visited a health facility or qualified provider, given the mean **quality** score | Proportion of women ages 15-49 with a live birth in the last two years who received ANC from a health facility, scaled by the mean ANC/nutrition quality score (i.e., provision/experience of care) of the stratum<br>**Note:** Provider quality (i.e., provision/experience of care) was calculated on a scale of [0,1] with 1 being perfect quality and 0 being no quality. Quality of care was defined for each intervention, accounting for whether the patient received the intervention or service according to clinical standards (including, e.g., appropriate history taking, assessment, counselling, use of diagnostics and treatment if appropriate) and with a positive experience. A mean provision/experience of care score was calculated for each stratum (i.e., category of qualified provider or health facility in each region/district). Please see the specific set of quality items in Error! Reference source not found. and Error! Reference source not found. for ANC and maternal nutrition. | HHS and HFA (direct observation) |
| User adherence-adjusted coverage | Proportion of the target population receiving the intervention(s) or service according to recommended standards and adhering to the treatment guidelines | Not estimated<br>**Note:** Adherence is not relevant for all interventions; in cases where the intervention is entirely delivered by the provider and there is no home care, behavior change, or continuing treatment needed, this component can be omitted. This is also difficult to define for a package of services such as ANC. | Not applicable |
| Outcome-adjusted coverage | Proportion of the target population experiencing the health gains from the service | Not estimated | Not applicable |

**Notes:** ANC = antenatal care, HFA = health facility assessment, HHS = household survey, IFA = iron and folic acid supplementation, SP = sulfadoxine-pyrimethamine, TT = tetanus toxoid vaccination

In 2016, WHO released new guidelines on ANC for a positive pregnancy experience, which recommends a minimum of eight ANC contacts (ANC8) as opposed to the earlier focused ANC model with its four recommended visits (ANC4) [16,17,30]. Due to the nature of multiple contacts with the health care system required for ANC, and variability in the expected timing of sequential visits, we have chosen to present three separate ANC cascades, defining service contact by the number of ANC visits (i.e., ANC1, ANC4+, ANC8+). We present the ANC8 cascade for all countries to reflect the implications of the new policy, although the majority of HHSs included in this analysis were conducted prior to 2016.

For this analysis, we explored both the complete package of services delivered through the ANC platform and the maternal nutrition interventions that are delivered during pregnancy. Although nutrition interventions are often delivered through a multi-faceted, multi-sectoral approach, antenatal consultations are an important platform for the delivery of nutrition interventions in pregnancy, especially as ANC attendance increases in LMICs [31,32]. In addition, HHSs and HFAs primarily collect data on maternal nutrition interventions delivered through ANC. As such, for the maternal nutrition cascade analysis, we focused on nutrition interventions delivered through the ANC platform.

### Country selection

The seven countries included in this analysis are Bangladesh, Haiti, Malawi, Nepal, Senegal, Sierra Leone, and Tanzania. We selected countries based on regional geographic diversity (representation from East Africa, Latin American and the Caribbean, South Asia, and West Africa) and on the availability of both HHS and HFA data. Ideally the HFA would have occurred within the two years before the HHS, and we would limit the analysis of household survey respondents to those with births in the last two years. We selected this reference period for several reasons. First, the two-year reference period is aligned with the Multiple Indicator Cluster Survey (MICS) and the Demographic and Health Surveys (DHS) version VIII approaches, which acknowledge the limitations of a long recall period [33-35]. While the DHS-7 asks questions related to ANC for births in the last five years, the DHS-8 asks about births in the last three years and restricts the analysis to births in the last two years. Second, for effective coverage estimation, if a five-year reference period were selected, the HHS data for births that occurred over a five-year period would be linked to HFA data representative of a period of a few months. To reduce the potential mismatch between the HHS and HFA reference periods, the HHS reference period was limited to two years. This allows for a sufficient sample size of women while better reflecting the potential care received based on the HFA data. The exception is Haiti, which has a three-year gap between the HHS and HFA. We relaxed our inclusion criteria for this country in order to include representation of Latin America and the Caribbean as no surveys met the two-year criteria.

At the time of data analysis, we found that very few countries had recent HFAs that measured the quality of service provision. Even fewer had a recent HFA and HHS that could be linked (i.e., where the HFA fell within the reference period for the HHS) [27]. As a result, we included two countries with no ANC service quality data (i.e., no direct observation of care) - Bangladesh and Sierra Leone. Thus, the endpoint for the coverage cascade for these two countries was intervention coverage.

### Data sources

For HHS data, we utilized data from the DHS program [36]. For the HFA data, we utilized data from the Service Provision Assessment (SPA) and the Service Availability and Readiness Assessment (SARA) [37,38]. The DHS and SPA are publicly available data. We obtained permission from the Sierra Leone Ministry of Health to use the 2017 SARA data. **Table 2** presents the countries and data sources included in the analysis.

**Table 2:** Countries and data sources included in the analysis.

| Country | HHS | HFA |
| --- | --- | --- |
| Bangladesh | DHS-VII 2014 | SPA 2014<sup>§</sup> |
| Haiti | DHS-VII 2016-17 | SPA 2013 |
| Malawi | DHS-VII 2015-16 | SPA 2013-14 |
| Nepal | DHS-VII 2016 | SPA 2015 |
| Senegal | DHS-VII 2017 | SPA 2016 |
| Sierra Leone | DHS-VI 2019 | SARA 2017<sup>§</sup> |
| Tanzania | DHS-VII 2015-16 | SPA 2014-15 |

§ These HFAs only include a facility audit component. No ANC client observation or ANC client exit interview was conducted.

### Household survey data

Data on care-seeking and intervention coverage among women with recent births were obtained from DHS. DHS are nationally representative HHSs with a large sample size that collect data on a wide range of population, health, and nutrition indicators. DHS survey data [33] was chosen over other available HHS data such as the MICS [35] because DHS collects information on the source of ANC care, which is needed for appropriate linkage of the HHS and HFA datasets [27]. Information on each HHS selected for this analysis can be found in Error! Reference source not found..

### Health facility assessment data

Data on health facilities and services were obtained from SPA and SARA surveys, which are HFAs that have many similarities and a few key differences [39]. SPA and SARA surveys both employ either a census or sample survey approach. Both surveys collect data on facility readiness through an inventory questionnaire, and the SPA and SARA programs have harmonized a core set of service readiness indicators that can be collected with either tool. SPA surveys include additional modules including a provider interview, client observations for ANC visits, and exit interviews of ANC clients. However, these modules are not implemented in every country. Information on each HFA selected for this analysis can be found in Error! Reference source not found..

While the SPA and SARA surveys are both HFAs, there are differences both between these surveys and within a survey program across countries. First, unlike SPA, the SARA does not capture any information on provision/experience of care. In addition, certain SPAs (e.g., Bangladesh 2014) did not include observation/exit interview modules and therefore did not collect provision/experience of care data. There are also differences in the readiness items captured, and in the service areas in which readiness items are recorded. Some readiness items are captured only in the SPA, not the SARA (e.g., examination bed). The SPA assesses the presence of equipment and infection prevention and control items in multiple service areas while the SARA focuses solely on the outpatient department. In addition, the SPA utilizes a staff roster to collect information on training for individual health workers while the SARA asks the service area in-charge whether at least one staff member providing the service has had training in the last two years. The SARA also focuses on broad areas of training (e.g., ANC training) while the SPA captures both broad training topics as well as more specific training areas (e.g., ANC screening, counseling for ANC, nutritional assessment of the pregnant woman). The SPA also has notable differences across countries. Some items are country-specific and therefore not collected in all SPAs (e.g., stadiometer only collected in Tanzania and Malawi). Finally, there are some services which are not part of the basic package of essential services in various countries due to the disease burden (e.g., HIV in Bangladesh; malaria in Bangladesh, Haiti, Nepal). As a result, these countries don’t include items related to those services (e.g., HIV diagnostic capacity, SP tablets, guidelines for IPTp). Details on HFA data availability are in Error! Reference source not found. and Error! Reference source not found..

### Defining and estimating coverage cascade steps

Error! Reference source not found. provides an overview of the indicators and data sources used to define each step of the coverage cascade for both ANC and maternal nutrition services delivered through ANC.

#### Target population

The target population, those in need of a service, was defined in the same way for both ANC and maternal nutrition cascades and was based on HHS data: women ages 15-49 years of age with a live birth in the last two years. The target population is the starting point for the effective coverage cascade, reflecting the population in need of the service or intervention which serves as the denominator for the cascade, and is therefore set at 100%.

#### Defining/estimating service contact

Service contact was defined in the same way for both ANC and maternal nutrition using three indicators and calculated from the HHS data: the proportion of women ages 15-49 with a live birth in the last two years who had at least one ANC contact (ANC1), four or more ANC contacts (ANC4), and eight or more ANC contacts (ANC8) during the most recent pregnancy. In determining the number of ANC contacts a woman received, women who reported receiving ANC but who were unsure of the number of contacts were assumed to have had at least one contact and therefore imputed one contact.

#### Defining/estimating intervention coverage

Intervention coverage was defined as the proportion of women 15-49 years of age with a live birth in the last two years who reported receiving key interventions (average score of interventions received) during the most recent pregnancy (**Table 1**), where the key interventions were defined separately for ANC and maternal nutrition. Similar to the content-qualified ANC coverage indicator (ANCq) approach proposed by Arroyave et al, intervention coverage estimates were generated at national level for each country by taking the average number of interventions received by each woman and then calculating the mean number of interventions across all women [40]. Indicator definitions for each of the key interventions are in Error! Reference source not found.. Not all key interventions were collected in all countries: Bangladesh excluded TT, SP for IPTp, IFA, and deworming; and Haiti and Nepal excluded SP for IPTp.

#### Defining/estimating readiness-adjusted and quality-adjusted coverage

##### Defining facility readiness and provision/experience of care (Item selection)

Readiness-adjusted coverage was defined as the proportion of women ages 15-49 with a live birth in the last two years who received ANC from a health facility, scaled by the mean ANC/nutrition readiness score of the stratum, where ANC readiness and nutrition readiness were defined separately for ANC and for maternal nutrition. Quality-adjusted coverage was defined similarly as the proportion of women ages 15-49 with a live birth in the last two years who received ANC from a health facility, scaled by the mean ANC/nutrition quality score (i.e., provision/experience of care) of the stratum, where ANC quality and maternal nutrition quality during ANC visits were defined separately for ANC and for maternal nutrition. To calculate readiness-adjusted coverage and quality-adjusted coverage, we first defined facility readiness and provision/experience of care for both ANC and maternal nutrition.

Selection of ANC and nutrition readiness and provision and experience of care items was guided by WHO guidelines and recommendations [16,17,30], expert surveys conducted by Sheffel et al [41] and King et al [42], and data availability based on selected HFA questionnaires [38,43]. The ANC and maternal nutrition readiness and provision/experience of care indices were developed independently with independent expert surveys, prioritization, and index development processes. This resulted in a few differences in intervention inclusion (e.g., the maternal nutrition index includes IPTp and deworming, while the ANC index does not) and more substantial differences in readiness and provision of care items included for each intervention (see Error! Reference source not found. and Error! Reference source not found. for details on item inclusion). We restricted the provision of care items to those required for a first ANC visit because for subsequent visits, the appropriate provision of care items are contingent on care received at previous visits and that information is not available in a cross-sectional facility survey. We also restricted the readiness analysis to items required for a first ANC visit to align with the selected provision of care items.

##### Calculating readiness and provision/experience of care scores (Index creation)

Because most of the readiness items were collected in the ANC module of the HFA questionnaire, we limited our analysis to facilities offering ANC services. For each facility or client included in the analysis, we defined binary (0/1) indicators for each item indicating whether the item was available at the time of the HFA. Binary indicators were created for all readiness items except for training, which was calculated from the health provider sections of the HFAs. Training indicators were defined as the proportion of health providers providing ANC services that had been trained in a particular intervention. A simple additive approach was utilized to generate an overall readiness score (unweighted average of the service readiness items available) and an overall provision/experience of care score (unweighted average of the provision/experience of care items received by a woman) for ANC and for maternal nutrition. For both ANC and maternal nutrition readiness indices and provision/experience of care indices, we found that items were relatively evenly distributed across domains and therefore the simple additive approach was appropriate for combining items into a single index score. This approach is consistent with a study on ANC which found small differences in the quality scores produced by three different approaches to combining items [41] as well as other studies which have taken a similar approach for developing quality of care indices for maternal and child health [42,44-46]. For the provision/experience of care index, we averaged all ANC client observations within a stratum (i.e., category of qualified provider or health facility in each region/district) rather than averaging ANC clients within individual facilities because we wanted to upweight higher caseload facilities within a stratum so that averages were representative of patient experience rather than representative of facilities. Averaging observations within a stratum implicitly weights the provision/experience of care indices by caseload (as caseload is correlated to number of observations in an HFA [47]).

##### Linking HHS and HFA data to estimate readiness-adjusted and quality-adjusted coverage

We used an ecological approach, which has been validated in three previous studies [48-50], to link each woman 15-49 years of age with a live birth in the last two years in the HHS to stratum-specific overall readiness scores and provision/experience of care scores calculated from the HFA.

We first mapped health facility categories (level and managing authority) in the HFA (SARA/SPA) to response options for place of ANC in the HHS (DHS). The HHS response options for place of ANC sometimes differed substantially from the HFA facility type categories. Where it was unclear how these aligned, we consulted with colleagues with country-specific expertise. In addition, not all qualified sources of care reported in the HHS were represented in the HFA, particularly for community health workers (CHWs) and mobile clinics. For each HHS, we reviewed the HHS questionnaire along with country-specific guidelines to determine which qualified providers were not captured in the HFA, and to assess the most appropriate approach for mapping these providers to facility types.

We then assigned each woman 15-49 years of age with a live birth in the last two years in the HHS to a stratum based on the place of ANC services (health facility type/managing authority) and place of residence (region in BGD, HTI, NPL, SEN, and TZA; district in MWI and SLE). Average readiness and provision/experience of care scores were calculated for each stratum of health facilities in the HFA. These scores ranged from 0 (no required items present) to 1 (all required items present). Each ANC care-seeking episode in the HHS was then assigned stratum average readiness and provision/experience of care scores based on the type of facility the woman reported attending for ANC. If a woman sought care from more than one provider, we took the average of the scores for the provider/facility types recorded and assigned that value to the woman (care-seeking from non-qualified sources was ignored). For example, a woman who received ANC at a public health center in Simiyu region, Tanzania, would be assigned the average readiness score for all public health centers in Simiyu region and the average provision/experience of care score for all women receiving ANC at public health centers in Simiyu region. In cases where no data was collected in the HFA for a facility type in a particular administrative area, the national average readiness score and provision/experience of care score for the corresponding health facility type was assigned to that stratum. Women who received ANC solely from unqualified providers were assigned a readiness score and provision/experience of care score of zero.

To estimate readiness-adjusted and quality-adjusted coverage in each country, we multiplied the binary ANC service contact coverage indicator (0/1) by the readiness score for each woman and the provision/experience of care score for each woman, respectively. We then calculated readiness-adjusted and quality-adjusted coverage at national level by taking an average of these values.

##### Analysis

We assessed each service contact coverage and intervention coverage indicator in each country for missing data and found that only Sierra Leone 2019 had missingness greater than 5% for IFA (for the number of times IFA was taken during the pregnancy). All service contact coverage, intervention coverage, and readiness- and quality-adjusted coverage estimates were generated using a design-based analysis to account for survey design. The mean readiness and provision of care scores were weighted using the SPA-calculated facility and client weights, respectively. The readiness-adjusted coverage and quality-adjusted coverage estimates were weighted using the DHS-calculated women’s weights. We used Taylor linearization to estimate the variance for service contact coverage and intervention coverage accounting for clustering and stratification in the HHSs. For readiness- and quality-adjusted coverage, we used a jackknife approach to account for the variance from both HHSs and HFAs, where the variance was derived from the distribution generated by withholding each household cluster and each health facility and re-estimating readiness- and quality-adjusted coverage [51]. All analyses were completed using R 4.1.3 [52], and R Studio[53]. To assist others who would like to implement the methods and/or replicate these results, the statistical code written for these analyses is publicly available at: <u>10.5281/zenodo.7671806</u>.

##### Ethical approval

This was a secondary analysis of publicly available, de-identified datasets and as such did not require ethical approval.

## Data Availability

All data produced in the present study are available upon reasonable request to the authors. To assist others who would like to implement the methods and/or replicate these results, the statistical code written for these analyses is publicly available.

https://zenodo.org/doi/10.5281/zenodo.7671805

## FINDINGS

### Background characteristics of surveys

The total analytical sample included interviews from 27,887 women with a birth in the last two years, audits from 7,523health facilities of which 6,441 provided ANC services, and direct observation of 4,261 ANC first visits across seven countries. Details of the sample are listed in **Table 3**.

**Table 3:** Total number of women, facilities, and ANC clients, by country.

| Country | Year | Number of women with a birth in the last two years | Total number of facilities | Number of facilities offering ANC | Number of ANC first visit clients |
| --- | --- | --- | --- | --- | --- |
| Bangladesh | DHS 2014, SPA 2014 | 3,217 | 1,596 | 1,493 | NA<sup>®</sup> |
| Haiti | DHS 2016-17, SPA 2013 | 2,583 | 907 | 832 | 785 |
| Malawi | DHS 2015-16, SPA 2013-14 | 6,814 | 1,060 | 643 | 859 |
| Nepal | DHS 2016, SPA 2015 | 2,007 | 992 | 872 | 573 |
| Senegal | DHS 2017, SPA 2016 | 4,926 | 484 | 323 | 307 |
| Sierra Leone | DHS 2019, SARA 2017 | 4,057 | 1,284 | 1,247 | NA<sup>®</sup> |
| Tanzania | DHS 2015-16, SPA 2014-15 | 4,283 | 1,200 | 1,031 | 1,737 |
| **Total** |  | **27,887** | **7,523** | **6,441** | **4,261** |

<sup>®</sup> The number of ANC first visit clients is not applicable (NA) in Bangladesh and Sierra Leone as these surveys did not include direct observation of care or client exit interviews.

### Source of care

**Figure 2** shows the sources of ANC within countries and across countries. Public primary-level facilities were the most often utilized source of care in every country except Bangladesh, where private hospitals were the most common source of ANC. Public hospitals were also an important source ANC in Haiti, Nepal, and Sierra Leone, where over one-third of women also sought care from a public hospital. Use of private facilities for ANC was low in all countries except Bangladesh where 50% of women sought care from a private hospital. Unqualified sources of care were utilized by less than 6% of women in all countries.

**Figure 2:**
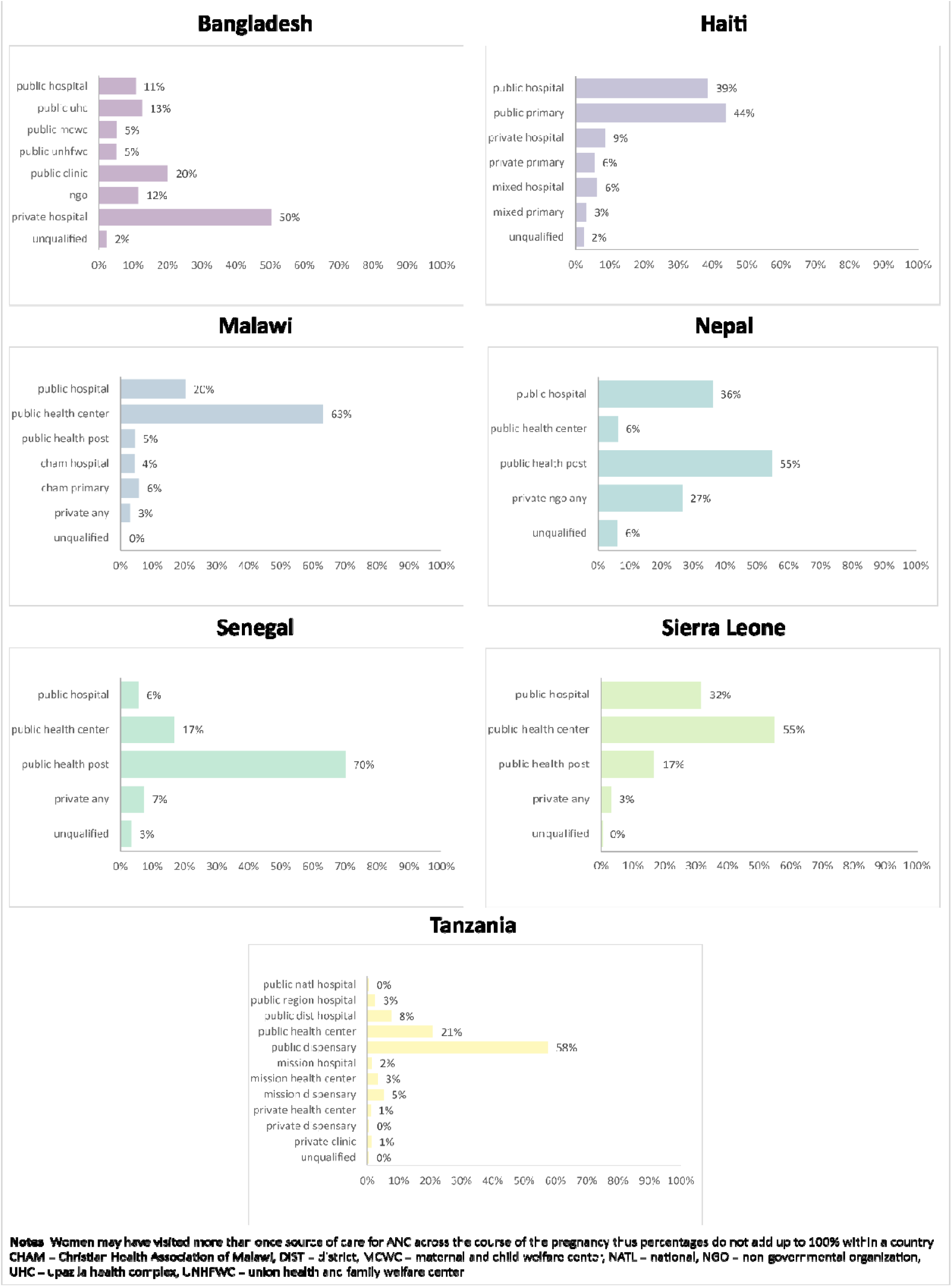
Variation in sources of ANC, by country.

### Readiness and provision/experience of care

#### Readiness

In general, ANC readiness decreased by level of facility within a managing authority (Error! Reference source not found.). For example, in Haiti, in the public sector there was a 20-point gap in average ANC readiness between hospitals and primary health facilities, with a similar pattern for the private sector and mixed facilities (26-point and 20-point gaps, respectively, in ANC readiness between hospitals and primary health facilities). Facilities of the same level but different managing authorities tended to have similar levels of readiness. There was a similar pattern for maternal nutrition readiness (Error! Reference source not found.), although the gaps were somewhat smaller than for ANC.

Individual ANC and maternal nutrition facility readiness items grouped by domains are presented by country in **Figure** *3* and **Figure 4** as well as Error! Reference source not found. and Error! Reference source not found.. Across countries, for both ANC and maternal nutrition, the domains of diagnostics and human resources had the lowest availability. For ANC, medicines and equipment tended to have higher availability while for maternal nutrition, equipment and basic amenities tended to have higher availability. Within the human resources domain, countries tended to perform better on the availability of general ANC guidelines and staff trained broadly in ANC but scored poorly on nutrition topic-specific guidelines and staff training. Individual item availability for ANC readiness was variable with some items being nearly universally available, such as iron and folic acid and adult weighing scale, while other items had more limited availability, such as hemoglobin testing capacity and staff trained in ANC screening and ANC counseling.

**Figure 3:**
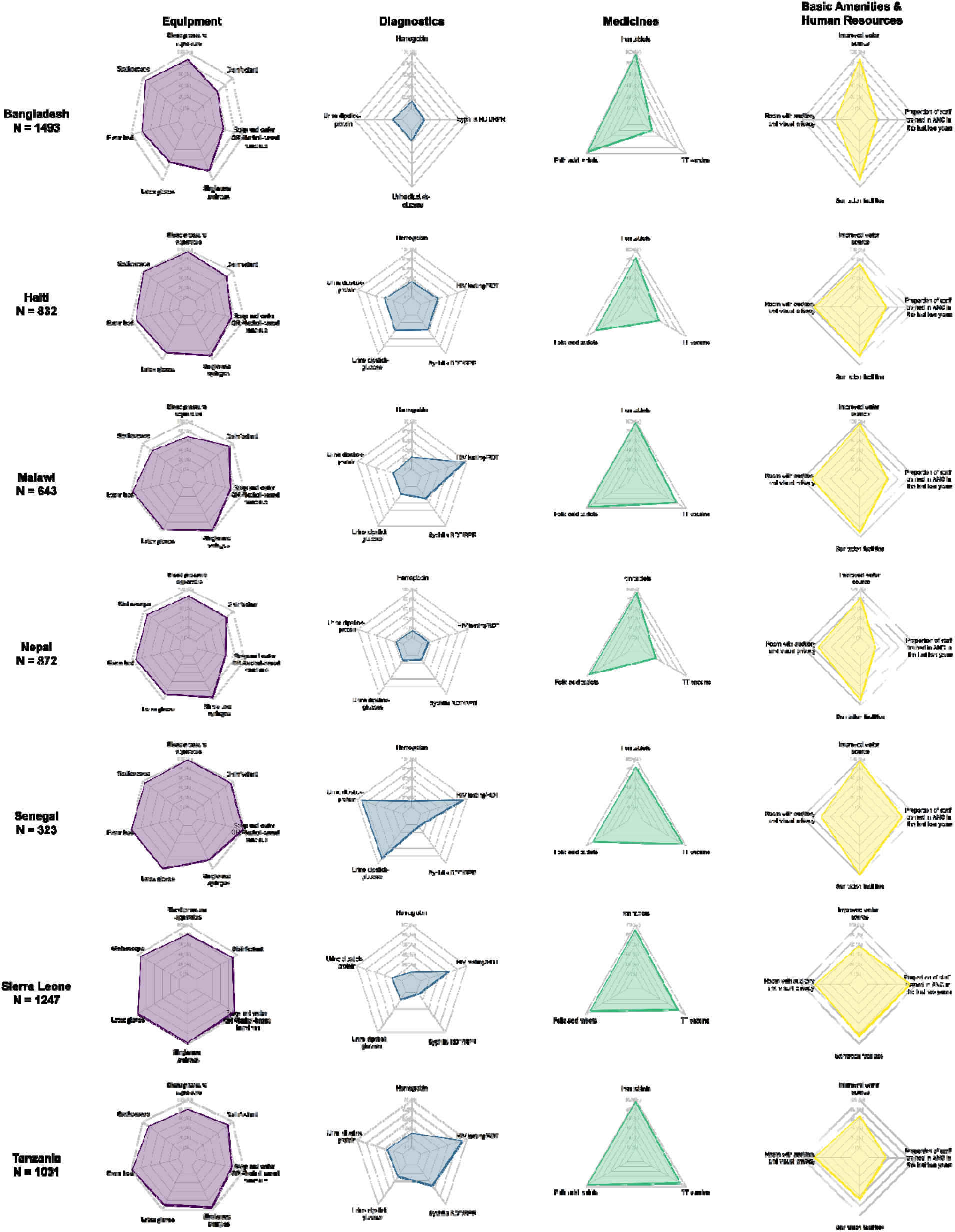
Availability of ANC readiness items, by domain and country.

**Figure 4:**
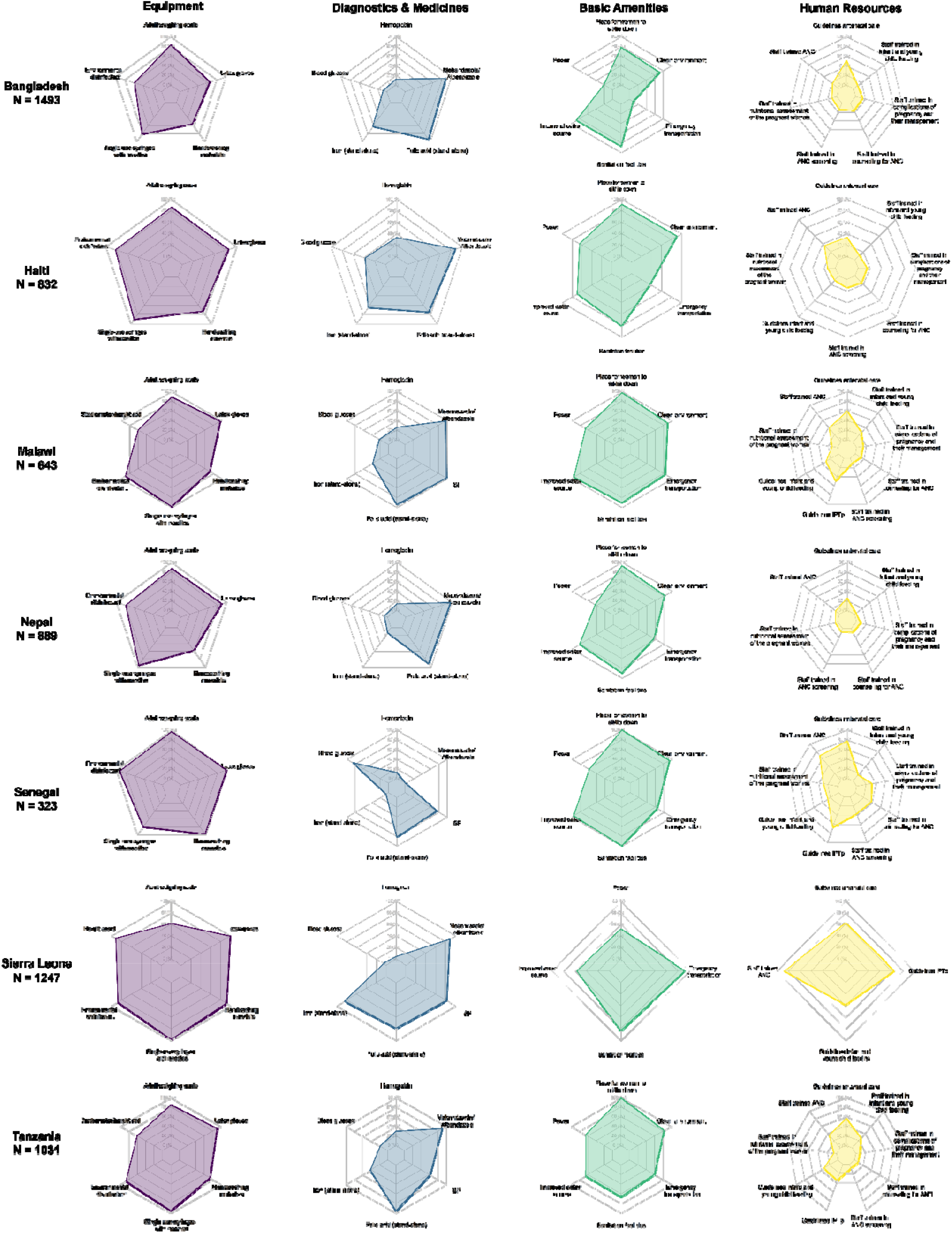
Availability of maternal nutrition readiness items, by domain and country.

#### Provision/experience of care

Across countries for both ANC and maternal nutrition, we found that providers at all levels of facilities across managing authorities provided services with a similar level of quality, and there was little variability within a country (Error! Reference source not found. and Error! Reference source not found.). Individual ANC and maternal nutrition provision/experience of care items by domain are presented by country in **Figure 5** and **Figure 6** as well as Error! Reference source not found. and Error! Reference source not found.. Across countries, providers were least likely to perform history-taking for complaints in pregnancy and client education and counseling while providers were more likely to perform observation and clinical investigation. In addition, clients reported a high level of satisfaction with the experience of care received. Some aspects of clinical care, such as weight and blood pressure assessment and provision of iron and/or folic acid, were almost universal, while syphilis testing, history-taking for a previous infant death, counseling on diet and nutrition, and counseling on exclusive breastfeeding were more limited.

**Figure 5:**
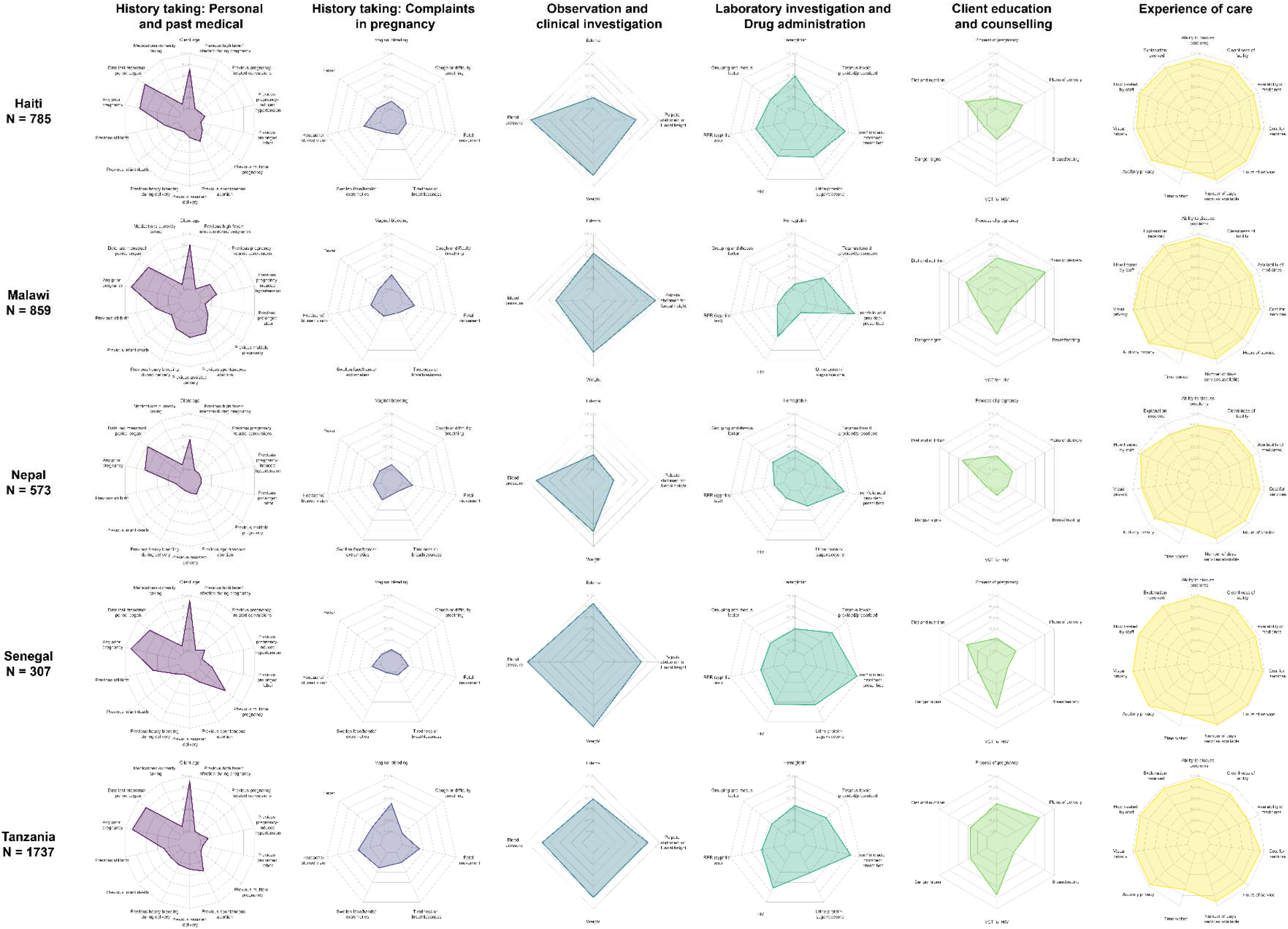
ANC provision/experience of care items, by domain and country.

**Figure 6:**
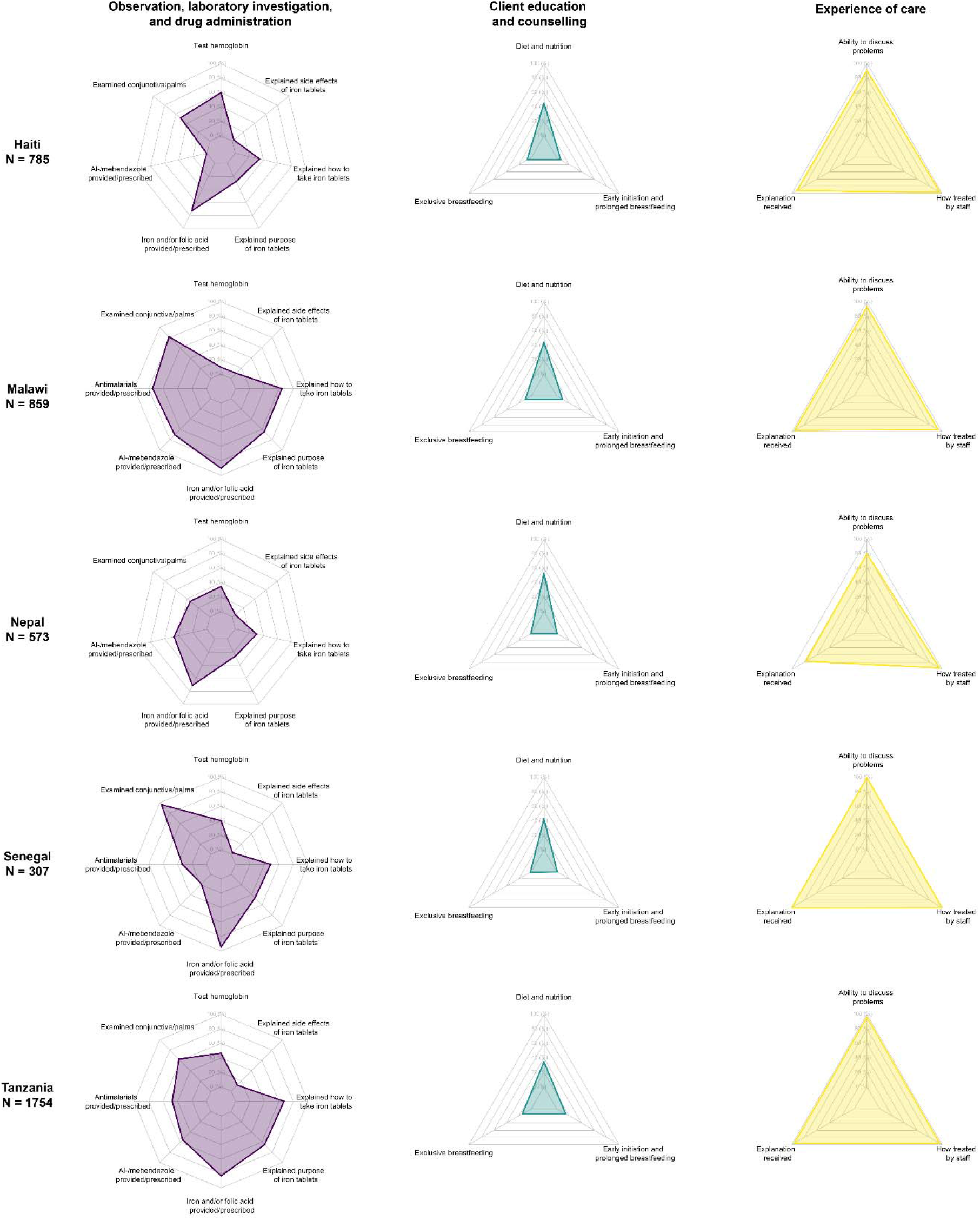
Maternal nutrition provision/experience of care items, by domain and country.

### Effective coverage cascades

#### ANC

The ANC effective coverage cascade estimates for the seven countries are presented in **Figure 7**, Error! Reference source not found., and Error! Reference source not found.. For the ANC1 cascade, as expected, coverage generally decreased across the cascade from service contact to quality-adjusted coverage. Service contact for at least one ANC visit was high across countries ranging from 80% in Bangladesh to 99% in Sierra Leone indicating high access to ANC services. The drop in coverage from service contact to readiness-adjusted coverage ranged from 20 percentage points (pp) in Senegal to 33 pp in Sierra Leone, highlighting a gap in ANC service readiness across countries. Intervention coverage was not consistently lower than readiness-adjusted coverage, indicating that some women reported receiving ANC services despite the level of service readiness at facilities. In Nepal and Sierra Leone, intervention coverage was 9 pp and 14 pp higher than readiness-adjusted coverage respectively. In the other countries, intervention coverage was 3-13 pp lower than readiness-adjusted coverage and represents a missed opportunity to deliver ANC services. Finally, quality-adjusted coverage was the lowest in all countries with the gap from intervention coverage to quality-adjusted coverage, representing inadequate service process, ranging from 7 pp in Tanzania to 44 pp in Nepal. Across the cascade, there was an average net decline in coverage of 52 pp (55% relative decrease) from service contact to quality-adjusted coverage, while the average net decline from service contact to intervention coverage was 28 pp (29% relative decrease).

**Figure 7:**
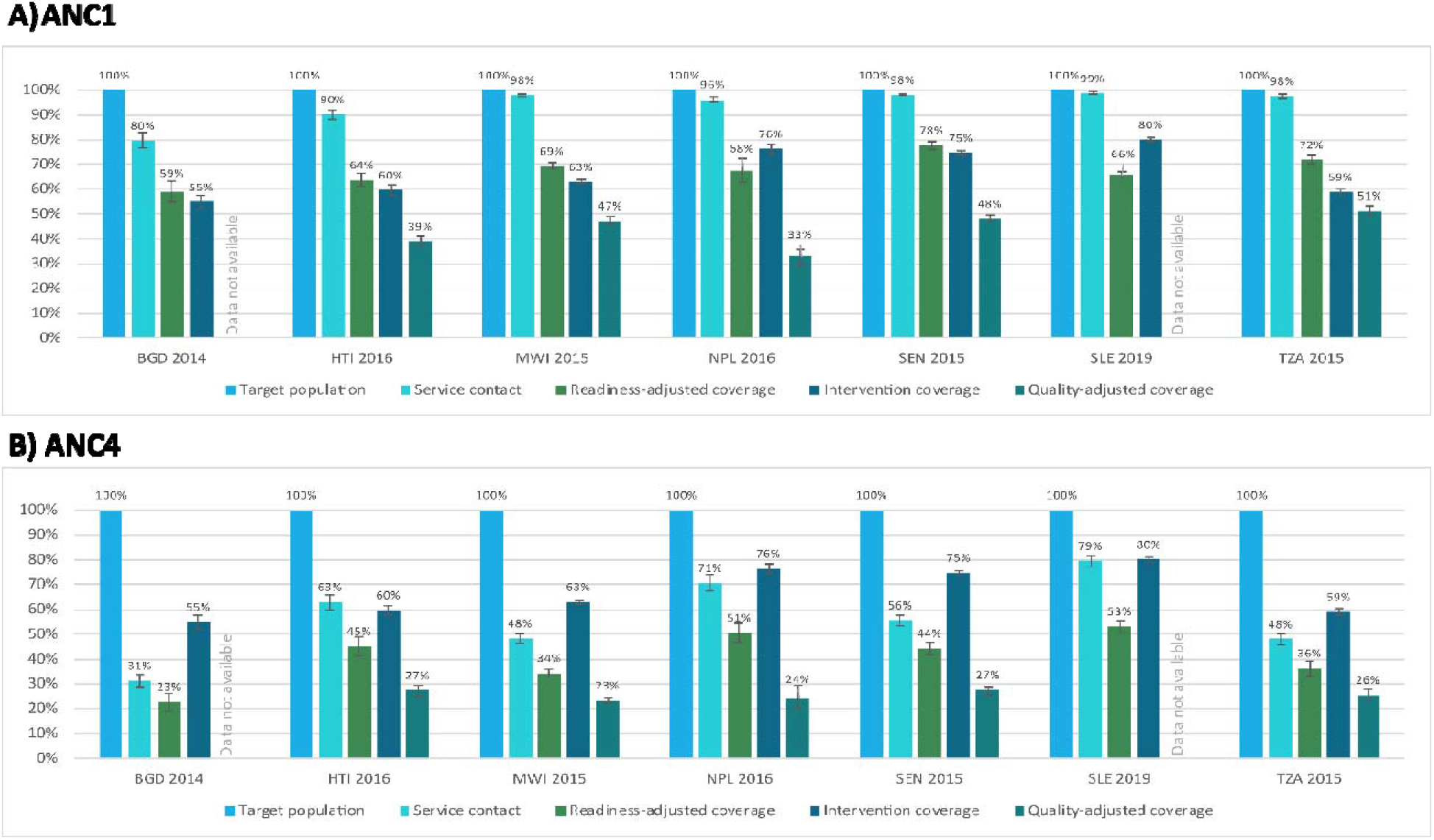
ANC effective coverage cascades, by country.

**Figure 8:**
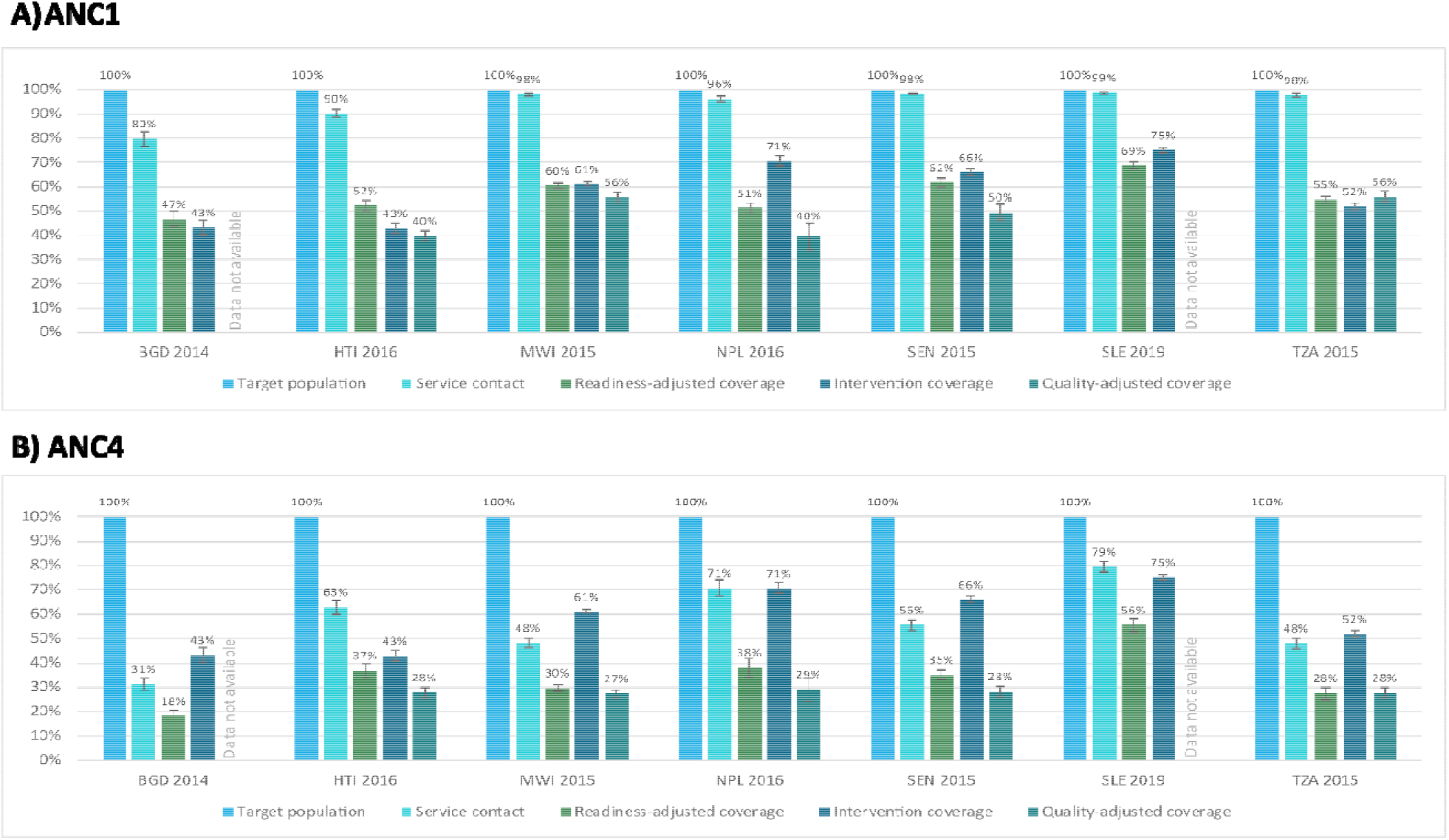
Maternal nutrition effective coverage cascades, by country.

The ANC4 and ANC8 coverage cascades followed a similar pattern as the ANC1 coverage cascades but with a few key differences. First, service contact was substantially lower in the ANC4 and ANC8 cascades, indicating lower access to four or more ANC visits. The proportion of women receiving at least four ANC visits ranged from 31% in Bangladesh to 79% in Sierra Leone while the proportion of women receiving at least eight ANC visits ranged from 0% in Senegal to 22% in Sierra Leone. The steep drop from target population to service contact in the ANC8 cascade left little room for further declines in readiness-adjusted and quality-adjusted coverage: on average, the net decline in coverage across the ANC8 cascade from service contact to quality-adjusted coverage was 3 pp (54% relative decrease). Second, intervention coverage was higher than readiness- and quality-adjusted coverage, because it was not restricted to women with 4+ or 8+ ANC visits. As a result, on average, the net increase in coverage across the cascade from service contact to intervention coverage was 10 pp for the ANC4 cascade (24% relative increase) and 59 pp for the ANC8 cascade.

#### Maternal nutrition

The maternal nutrition effective coverage cascade estimates for the seven countries are presented in

**Figure** 8, Error! Reference source not found., and Error! Reference source not found.. Comparisons between the ANC and maternal nutrition quality-adjusted and readiness-adjusted estimates are in Error! Reference source not found.. The first two steps of the maternal nutrition cascades – target population and service contact – were the same as for the ANC cascades.

For the ANC1 maternal nutrition cascade, the drop in coverage from service contact to readiness-adjusted coverage ranged from 30 pp in Sierra Leone to 45 pp in Nepal, reflecting a larger gap in service readiness for nutrition interventions than for general ANC interventions. Similar to the overall ANC cascades, intervention coverage was not consistently lower than readiness-adjusted coverage, indicating that some women reported receiving maternal nutrition services despite low levels of service readiness at facilities. Quality-adjusted coverage for maternal nutrition was slightly higher than quality-adjusted coverage for ANC in all countries. On average there was a net decline in coverage of 48 pp (50% relative difference) across the ANC1 maternal nutrition cascade from service contact to quality-adjusted coverage and 38 pp (38% relative difference) from service contact to intervention coverage, similar to the gap in the overall ANC1 cascades. The ANC4 and ANC8 maternal nutrition coverage cascades were similar to the ANC1 maternal nutrition coverage cascades. The key differences between the ANC4 and ANC8 maternal nutrition coverage cascades and ANC1 maternal nutrition cascades mirror those found for the ANC cascades.

Our effective coverage cascade operationalization for ANC and maternal nutrition services provides a rigorous approach to estimating effective coverage for these services by linking data sources commonly available in LMICs. Applying a set of best practices for effective coverage estimation, we found that effective coverage cascades provided a summary measure of service coverage and quality [27]. While the aim of this work was to identify the challenges in operationalizing these cascades rather than to estimate “effective coverage” for a particular country, we found that across countries and service areas we generally saw a substantial drop from service contact to readiness-adjusted coverage and a further drop to quality-adjusted coverage. Our findings add to a wealth of evidence demonstrating large drops in coverage once adjustments for service quality (readiness and process dimensions) are accounted for [23], thus underscoring the critical role of service quality in producing better health outcomes [7,54]. However, we encountered challenges in operationalizing the effective coverage cascades, including limited data availability, standardization and comparability, and methodological complexity.

### Added value of effective coverage cascades

Our analyses provide an example of how the coverage cascade approach can highlight gaps in service access, service readiness, and quality of care, allowing for tailored interventions to address setting-specific challenges in the delivery of health services [22,23]. This approach to assessing implementation bottlenecks in health service delivery was strengthened by a detailed analysis of facility readiness and provision/experience of care which provided for more actionable recommendations on how to strengthen health systems. In addition, estimating readiness- and quality-adjusted coverage accounted for not only the overall level of quality at facilities, but also the level of quality in the types of facilities at which women are most commonly accessing care.

Our operationalization of the ANC and maternal nutrition cascades shed light on which steps of the cascade can be estimated. We did not estimate step 5 or step 6, user- or outcome-adjusted coverage, for ANC or maternal nutrition. Some ANC interventions do not require user adherence (e.g., TT injection) and among those that do, limited data on user adherence is collected. For example, adherence is required for IFA and calcium supplementation during pregnancy. Recent studies have found poor validity of IFA supplementation coverage and adherence questions in HHSs [55], and there is no data on calcium supplementation coverage or adherence in DHS or MICS. Outcome-adjusted coverage is not the ideal measure of ANC effective coverage as health outcomes such as reduced maternal mortality, neonatal mortality, and stillbirth are more strongly associated with the quality of labor and delivery care than ANC [10,56,57]. As such, the endpoint of the ANC care cascade and the preferred measure of effective coverage for ANC is step four, quality-adjusted coverage [22].

#### Challenge 1: Limited data availability, particularly for maternal nutrition

Data availability proved to be a challenge for operationalizing the ANC and maternal nutrition effective coverage cascades. While the data for ANC is quite complete, we found more data gaps related to maternal nutrition, whereby some interventions were not represented in our analysis (e.g., calcium supplementation during pregnancy) [42]. In addition, recommended nutrition interventions provided during ANC are largely counselling-based, which is not captured well across data sources including HHSs and HFAs.

We also found discrepancies between data available in HHSs to estimate intervention coverage as compared to data available in HFAs to estimate readiness and provision of care. The measure of intervention coverage used for this analysis relies on reported receipt of seven interventions which pertain to observation and clinical investigation, laboratory investigation, and drug administration.

Including only these seven ANC interventions assumes that these items are a good tracer for other interventions comprising a high-quality ANC service. Of these seven interventions, two (tetanus toxoid and iron-folic acid supplementation) can be delivered through multiple channels and not necessarily only during ANC. This is particularly true of tetanus toxoid vaccination coverage which is based on lifetime doses, meaning that a woman and her baby can be protected without having received any TT doses during the most recent pregnancy. In addition, this set of interventions does not capture important aspects of an ANC visit including history taking, counselling, and the client experience of care, which are equally important and well detailed in ANC clinical care guidelines but are difficult to ask about in a HHS. Based on our analysis of SPA observation data, we also found that these elements are the service components most likely to be excluded from an ANC visit. As a result, the intervention coverage measure may not accurately reflect the service quality received by pregnant women. The DHS-8 has added in four additional validated measures of content of care to assess the content of care more comprehensively, including listening to the baby’s heartbeat, nutrition counseling, breastfeeding counseling, and discussion of pregnancy danger signs (i.e., vaginal bleeding) [34]. Future HHSs thus may have a more comprehensive measure of intervention coverage, although using these additional questions will create challenges with historical comparability and cross-country comparability (when some countries have an older DHS survey and others have a new DHS survey).

Our experience of care measures incorporate aspects of respectful care; however previous studies have noted methodological challenges such as courtesy bias resulting in universally high levels of client satisfaction [41,58]. Validated measures of respectful ANC are in the early stages of development [59-61] and a validated 8-item scale measuring person centered ANC has been incorporated into the 2022 SPA questionnaires which may improve data availability going forward [62].

Finally, data on clinical quality of ANC and maternal nutrition is only available in a limited number of countries – namely those that have implemented a SPA survey. Future survey implementation of the Harmonized Health Facility Assessment (the successor to the SARA) may provide measures of clinical quality through a record review approach [63]. However, innovative methods to measure process quality on a more routine basis are needed to ensure availability of data across countries and at regular intervals. Effective coverage cascades are thus not a panacea for data gaps, and in fact highlight the need to improve the availability of data across LMICs to ensure countries have the data they need to track the ability of their health systems to delivery high quality care to all in need and make decisions on priority investments or improved intervention implementation.

#### Challenge 2: Standardization and comparability

This analysis highlighted that effective coverage, while conceptually attractive, is quite complex and may prove challenging to implement on a larger scale while ensuring standardization and comparability of cascades between countries. The lack of standardized definitions and indicators for measuring maternal health service quality continues to pose a challenge to constructing effective coverage cascades, particularly for global monitoring or cross-country comparison [26,64,65]. While work has been done to propose indicators and indices of service readiness and quality of care for ANC and maternal nutrition, there remains no consensus on standardized indicators of maternal quality of care [41,42,66]. Within our own analysis, we found that HFA tools were not fully harmonized resulting in different data availability depending on the type of survey used for the analysis and country-specific tool adaptations. This has implications for trying to generate effective coverage estimates that are comparable over time or across countries. The cascades that we generated are not therefore directly comparable across countries but may still be useful for within country monitoring and service delivery improvement. We chose not to standardize items to only those available across countries as this may not be indicative of a “high quality” service in a particular country. While there was some variation in data availability across countries, data availability overall was relatively high as compared to other service areas, such as postnatal care and small & sick newborns [67,68]. ANC was one of the first service packages to be standardized with the focused ANC guidelines [16] which has enabled the development of tools designed to capture aspects of quality for the full package of interventions. In addition, as ANC is a preventative care platform, standardizing the required items and tools is more straightforward.

#### Challenge 3: Methodological complexity

Numerous methodological decisions must be made to generate effective coverage estimates. While we followed a standard approach as per Munos et al, there were additional considerations for ANC and maternal nutrition specifically [27]. First, women may have multiple ANC visits across the course of the pregnancy and therefore may seek care from multiple sources. In these cases, we chose to take the average readiness score from the multiple sources of care to reflect the average client experience. Alternative approaches include utilizing the source of care with the highest or lowest readiness which would result in maximum or minimum measures of service quality but might be less reflective of the client service delivery experience.

Second, this approach to effective coverage estimation does not capture non-facility-based interventions such as community-based care and informal sources of care such as pharmacies [27]. This is particularly a relevant issue for maternal nutrition as these interventions often have a community-based delivery platform in addition to facility-based care. Our measures of intervention coverage were sometimes higher than readiness-adjusted coverage; the receipt of ANC and maternal nutrition interventions outside of facility-based care that were not captured in this analysis may be a contributing factor to this finding.

Third, as the index development processes for ANC and maternal nutrition were independent and relied on independent groups of experts, in some cases an item was included in the nutrition readiness index but excluded in the ANC index (e.g., emergency transportation) [41,42]. We note that these decisions on what to include and exclude in the indices may lead to differences in the readiness and quality of care estimates and therefore the readiness-adjusted and quality-adjusted estimates. This can make it difficult to compare effective coverage across services and analyses and highlights the difficulty and importance of index development for use in effective coverage cascades.

Finally, utilizing a HFA which incorporates direct observation of care and client exit interviews to estimate quality-adjusted coverage generates a measure of service quality that more accurately reflects the true quality of ANC a woman received. Direct observation allows for capturing information on adherence to clinical guidelines during service delivery, and the client exit interview allows for incorporation of aspects of client experience. However, there are some limitations to using this data in an effective coverage analysis. Using an ecological approach to link HFA and HHS data does not provide information on what a specific woman got at a particular visit; individual health records would be needed for this type of analysis. In addition, if the timing of the HHS and HFA are not well aligned, the time lag may result in quality-adjusted coverage measures that may not accurately reflect the service quality received by pregnant women in the household survey. The additional validated ANC content of care measures added to the DHS-8 have improved the feasibility of measuring ANC service quality from a HHS, which may diminish the added value of quality-adjusted coverage for ANC given these methodological challenges [40]. However future research is needed to enhance our understanding of whether the “new” intervention coverage measures are good proxies for ANC service quality as measured through direct observations [40].

#### Limitations

Our study has some limitations. Because of data limitations, we used ecological linking of HHS and HFA datasets which means that women were not linked to the specific facility in which they sought care but rather a facility average for the type of facility in the region which they sought care. There are studies that suggest that this approach produces valid estimates, but there is a possibility of bias under certain conditions such as when there is a lot of variability in readiness and quality and/or high levels of preferential careseeking [49,69]. In addition, due to data limitations, we linked datasets with a two to three year time difference. However, we utilized the most robust methods available for the data that was available to us. These methodological choices and resulting limitations should be taken into consideration when deciding when and how to use effective coverage measures of ANC and maternal nutrition. In addition, this work relied on survey data and did not utilize routine health information system data for measures of service readiness or service quality. More research is needed to determine the availability and feasibility of using routine data for effective coverage estimation. Our readiness and quality measures were based on expert surveys, the WHO quality of care framework, and data availability and were not always comparable across countries. However, our approach to index development followed a rigorous process and was supported by evidence that overall ANC readiness and quality scores are not greatly affected by the addition or removal of a few items [41]. Finally, this study did not include country consultation on the item inclusion for readiness and quality indices, as the aim of this work was to identify the challenges in operationalizing these cascades rather than to estimate “effective coverage” for a particular country. However, this method’s application has shown that much country adaptation is required to make these analyses relevant to a given country context, and more work is needed to understand the utility of these measures for country stakeholders.

## CONCLUSIONS

The coverage cascade approach yielded summary measures that were useful for identifying high-level barriers to effective coverage of antenatal care and maternal nutrition; however, detailed measures within the cascade – such as source of care, readiness and quality scores by domain, or the availability of individual items – may be needed for evidence-based decision making. This exercise highlights how an effective coverage cascade may illustrate the potential bottlenecks in achieving expected health benefits from services and the inter-connectedness of service access and service quality in achieving improvements in health status. Increased collection and standardization of ANC and maternal nutrition quality of care data is needed to improve measurability and comparability of effective coverage cascades. Future work on usability of routine data along with analyses focusing on subregional effective coverage and disaggregation based on level of health facility may improve the usefulness of effective coverage estimates for targeting interventions and resource allocation.

## DECLARATIONS

## Acknowledgements

The authors wish to acknowledge the Bill & Melinda Gates Foundation for their support of this project. We are grateful to the Improving Measurement & Program Design (IMPROVE) core group for their technical inputs and expertise during the development of this analysis, in particular Ann Blanc, Allisyn Moran, and Cindy Stanton. We additionally would like to thank Malick Kante, Tsering Lama, and Meike Schleiff for their contributions to understanding provider cadres.

## Ethics approval

This is a secondary analysis and as such did not involve human subjects research.

## Authors’ contributions

AS, EC, and MKM contributed to conceptualizing the paper and analysis. AS conducted the analysis and prepared the manuscript. All authors critically reviewed and revised the manuscript and approved the final manuscript.

