## Supplementary_Material for "Effective coverage for maternal health: operationalizing effective coverage cascades for antenatal care and nutrition interventions for pregnant women in seven low- and middle-income countries"

**Supplementary Materials**

Supplementary Table 1: Household survey data sources

| **Country** | **HHS** | **Study design details** | **Sample size** |
| --- | --- | --- | --- |
| Bangladesh | DHS 2014 | Sample representative nationally as well as of urban and rural areas and division | 3,217 women with a birth in the last two years |
| Haiti | DHS 2016-17 | Sample representative nationally as well as of urban and rural areas and department | 2,583 women with a birth in the last two years |
| Malawi | DHS 2015-16 | Sample representative nationally as well as of urban and rural areas, region, and district | 6,814 women with a birth in the last two years |
| Nepal | DHS 2016 | Sample representative nationally as well as of urban and rural areas, development region, and province | 2,007 women with a birth in the last two years |
| Senegal | DHS 2017 | Sample representative nationally as well as of urban and rural areas and region | 4,926 women with a birth in the last two years |
| Sierra Leone | DHS 2019 | Sample representative nationally as well as of urban and rural areas, region, and district | 4,057 women with a birth in the last two years |
| Tanzania | DHS 2015-16 | Sample representative nationally as well as of urban and rural areas, zone, and region (for most indicators) | 4,283 women with a birth in the last two years |

Supplementary Table 2: Health facility assessment data sources

| **Country** | **HFA** | **Census or Sample** | **Study design details** | **Modules implemented** |
| --- | --- | --- | --- | --- |
| Bangladesh | SPA 2014 | Sample | Stratified random sample of 1,596 health facilities selected from all formal-sector health facilities in Bangladesh. Designed to be representative of the seven administrative divisions and by facility type. | Facility inventory |
| Haiti | SPA 2013 | Census | Census of all health facilities (public and private) with a sample of providers and clients at facilities. | Facility inventory, ANC client observation, ANC client exit interview |
| Malawi | SPA 2013-14 | Census | Census of all health facilities (public and private) with a sample of providers and clients at facilities. | Facility inventory, ANC client observation, ANC client exit interview |
| Nepal | SPA 2015 | Sample | Stratified random sample of 992 health facilities selected from all formal-sector health facilities in Nepal. Designed to be representative of the three geo-ecological regions in the country, the 13 development-ecological zones, by facility type, and by  managing authority. | Facility inventory, ANC client observation, ANC client exit interview |
| Senegal | SPA 2016 | Sample | Stratified random sample of 484 health facilities selected from all formal-sector health facilities in Senegal. Designed to be representative by facility type. | Facility inventory, ANC client observation, ANC client exit interview |
| Sierra Leone | SARA 2019 | Census | Census of all health facilities (public and private). | Facility inventory |
| Tanzania | SPA 2014-15 | Sample | Stratified random sample of 1,200 health facilities selected from all formal-sector health facilities in Tanzania. Designed to be representative by region, facility type, and managing authority. | Facility inventory, ANC client observation, ANC client exit interview |

Supplementary Table 3: ANC readiness and provision/experience of care items and detailed definitions

| **No** | **Domain** | **Code** | **Item** | **Detailed indicator definition** | **Data availability notes** | **Survey specific: Indicator not collected** | **Cross-country differences** |
| --- | --- | --- | --- | --- | --- | --- | --- |
| **Readiness** | | | | | | | |
| 1 | Equipment and supplies | ANC_READY_03 | Blood pressure apparatus | Either manual or digital blood pressure apparatus observed available and functional in the ANC service provision area. | SARA only asks about this item in the OPD |  | Sierra Leone is OPD instead of ANC service area |
| 2 | Equipment and supplies | ANC_READY_30 | Stethoscope | Stethoscope observed available and functional in the ANC service provision area. | SARA only asks about this item in the OPD |  | Sierra Leone is OPD instead of ANC service area |
| 3 | Equipment and supplies | ANC_READY_22 | Examination bed | Examination bed observed available in the ANC service provision area. | Not collected in SARA | SLE 2017 |  |
| 4 | Equipment and supplies | GEN_READY_17 | Latex gloves | Clean/sterile disposable latex gloves observed available in the OPD or ANC service provision area. | SARA only asks about this item in the OPD |  | Sierra Leone is OPD only |
| 5 | Equipment and supplies | GEN_READY_15 | Auto-disable syringes with needles; single use standard disposable syringes with needles | Single-use standard disposable syringes with needles or auto-disable syringes with needles observed available anywhere in the facility. | SARA only asks about this item in the OPD whereas SPA is in multiple service areas |  | Sierra Leone is OPD only |
| 6 | Equipment and supplies | GEN_READY_16 | Handwashing materials | Either handwashing soap and running water OR alcohol-based hand sanitizer observed available in the OPD or ANC service provision area. | SARA only asks about this item in the OPD |  | Sierra Leone is OPD only |
| 7 | Equipment and supplies | GEN_READY_14 | Environmental disinfectant | Disinfectant (e.g., chlorine, hibitane, alcohol) observed available anywhere in the facility. | SARA only asks about this item in the OPD whereas SPA is in multiple service areas |  | Sierra Leone is OPD only |
| 8 | Diagnostics | ANC_READY_04 | Hemoglobin | Facility has the ability to conduct hemoglobin testing onsite and has the required equipment available and functioning/unexpired (rapid test OR hematology analyzer OR hemocue and microcuvette OR colorimeter or hemoglobinometer & drabkin's solution & pipette OR litmus paper for hemoglobin test). |  |  | Haiti, Malawi, Nepal, Senegal, and Bangladesh don't include functioning of microcuvette or drabkin solution unexpired Nepal also doesn't include litmus paper for hemoglobin test Sierra Leone only includes heomcue, colorimeter, hemoglobinometer |
| 9 | Diagnostics | ANC_READY_05 | Urine dipstick- protein | Facility has the ability to conduct urine protein dipstick testing onsite and has the required equipment available and functioning/unexpired (dipsticks for urine protein) |  |  |  |
| 10 | Diagnostics | ANC_READY_23 | Urine dipstick- glucose | Facility has the ability to conduct urine glucose dipstick testing onsite and has the required equipment available and functioning/unexpired (dipsticks for urine glucose) |  |  |  |
| 11 | Diagnostics | ANC_READY_25 | Syphilis RDT / RPR | Facility has the ability to conduct syphilis RDT / RPR testing onsite and has the required equipment available and functioning/unexpired (RDT, PCR, VDLR, RPR, rotator/shaker). |  |  | Sierra Leone doesn't ask about rotator/shaker |
| 12 | Diagnostics | ANC_READY_26 | HIV diagnostic capacity | Facility has the ability to conduct HIV testing onsite and has the required equipment available and functioning/unexpired (RDT, ELISA (scanner/reader, washer, assay, and incubator), dynabeads, or Western blot ). |  | BGD 2014 | BGD does not assess HIV MWI doesn't ask about expiration of western blot or availability of ELISA assay or incubator MWI doesn't ask about expiration of western blot or availability of ELISA assay NPL doesn't ask about expiration of western blot, RDT in ANC service site, RDT in FP service site Sierra Leone doesn't ask about dynabeads or Western blot, RDT is only asked in one location |
| 13 | Medicines and commodities | ANC_READY_06 | Iron tablets | Iron tablets or iron + folic acid combination tablets observed in pharmacy or anywhere in the facility where medicines are routinely stored; at least one with valid expiration date. |  |  |  |
| 14 | Medicines and commodities | ANC_READY_07 | Folic acid tablets | Folic acid tablets or iron + folic acid combination tablets observed in pharmacy or anywhere in the facility where medicines are routinely stored; at least one with valid expiration date. |  |  |  |
| 15 | Medicines and commodities | ANC_READY_08 | Tetanus toxoid vaccine | Tetanus toxoid vaccine observed in pharmacy or anywhere in the facility where medicines are routinely stored; at least one with valid expiration date. |  |  |  |
| 16 | Basic amenities | GEN_READY_02 | Improved water source | Facility has an improved water source that is onsite or within 500meters of the facility. |  |  |  |
| 17 | Basic amenities | GEN_READY_03 | Room with auditory and visual privacy | Room is private room with auditory and visual privacy is observed in the OPD. |  |  |  |
| 18 | Basic amenities | GEN_READY_04 | Sanitation facilities | Facility has a functioning toilet (latrine) available for general outpatient client use. |  |  |  |
| 19 | Human resources | ANC_READY_02 | Staff trained in the past 2 years in antenatal care (broad ANC training) | Proportion of health care workers delivering ANC services that have been trained in antenatal care (regardless of the specific topic area) in the last two years.  Alternative: At least one health care worker providing ANC services has been trained in antenatal care (regardless of the specific topic area) in the last two years. |  |  | Sierra Leone uses alternative definition while SPA countries use preferred/ original definition |
| **Provision/experience of care** | | | | | | | |
| 20 | History taking | ANC_PROV_01 | Provider asked about client age | During the observed ANC visit, the provider took a personal history and asked about the client's age. | Not collected in SARA; Collected only in some SPAs (those that include client/provider observation) | BGD 2014 SPA SLE 2017 SARA |  |
| 21 | History taking | ANC_PROV_02 | Provider asked about medications client is taking | During the observed ANC visit, the provider took a personal history and asked about medications the client is taking. | Not collected in SARA; Collected only in some SPAs (those that include client/provider observation) | BGD 2014 SPA SLE 2017 SARA |  |
| 22 | History taking | ANC_PROV_03 | Provider asked about date last menstrual period began | During the observed ANC visit, the provider took a personal history and asked about the date the client's last menstrual period began. | Not collected in SARA; Collected only in some SPAs (those that include client/provider observation) | BGD 2014 SPA SLE 2017 SARA |  |
| 23 | History taking | ANC_PROV_04 | Provider asked about any prior pregnancy | During the observed ANC visit, the provider took a personal history and asked about any prior pregnancy. | Not collected in SARA; Collected only in some SPAs (those that include client/provider observation) | BGD 2014 SPA SLE 2017 SARA |  |
| 24 | History taking | ANC_PROV_06 | Provider asked about previous still birth | During the observed ANC visit, the provider took a medical history for prior pregnancies and asked about still birth. | Not collected in SARA; Collected only in some SPAs (those that include client/provider observation) | BGD 2014 SPA SLE 2017 SARA |  |
| 25 | History taking | ANC_PROV_07 | Provider asked about prior pregnancy in which the infant died in the first week of life | During the observed ANC visit, the provider took a medical history for prior pregnancies and asked about any infant that died in the first week of life. | Not collected in SARA; Collected only in some SPAs (those that include client/provider observation) | BGD 2014 SPA SLE 2017 SARA |  |
| 26 | History taking | ANC_PROV_08 | Provider asked about heavy bleeding during or after delivery in prior pregnancies | During the observed ANC visit, the provider took a medical history for prior pregnancies and asked about heavy bleeding during or after delivery. | Not collected in SARA; Collected only in some SPAs (those that include client/provider observation) | BGD 2014 SPA SLE 2017 SARA |  |
| 27 | History taking | ANC_PROV_09 | Provider asked about previous assisted delivery | During the observed ANC visit, the provider took a medical history for prior pregnancies and asked about previous assisted delivery. | Not collected in SARA; Collected only in some SPAs (those that include client/provider observation) | BGD 2014 SPA SLE 2017 SARA |  |
| 28 | History taking | ANC_PROV_10 | Provider asked about previous spontaneous abortion | During the observed ANC visit, the provider took a medical history for prior pregnancies and asked about previous spontaneous abortion. | Not collected in SARA; Collected only in some SPAs (those that include client/provider observation) | BGD 2014 SPA SLE 2017 SARA |  |
| 29 | History taking | ANC_PROV_11 | Provider asked about previous multiple pregnancies | During the observed ANC visit, the provider took a medical history for prior pregnancies and asked about previous multiple pregnancies | Not collected in SARA; Collected only in some SPAs (those that include client/provider observation) | BGD 2014 SPA SLE 2017 SARA |  |
| 30 | History taking | ANC_PROV_12 | Provider asked about previous prolonged labor | During the observed ANC visit, the provider took a medical history for prior pregnancies and asked about previous prolonged labor. | Not collected in SARA; Collected only in some SPAs (those that include client/provider observation) | BGD 2014 SPA SLE 2017 SARA |  |
| 31 | History taking | ANC_PROV_13 | Provider asked about previous pregnancy-induced hypertension | During the observed ANC visit, the provider took a medical history for prior pregnancies and asked about pregnancy-induced hypertension. | Not collected in SARA; Collected only in some SPAs (those that include client/provider observation) | BGD 2014 SPA SLE 2017 SARA |  |
| 32 | History taking | ANC_PROV_14 | Provider asked about previous pregnancy related convulsions | During the observed ANC visit, the provider took a medical history for prior pregnancies and asked about pregnancy related convulsions. | Not collected in SARA; Collected only in some SPAs (those that include client/provider observation) | BGD 2014 SPA SLE 2017 SARA |  |
| 33 | History taking | ANC_PROV_15 | Provider asked about high fever or infection during prior pregnancy/pregnancies | During the observed ANC visit, the provider took a medical history for prior pregnancies and asked about high fever or infection during a prior pregnancy/pregnancies. | Not collected in SARA; Collected only in some SPAs (those that include client/provider observation) | BGD 2014 SPA SLE 2017 SARA |  |
| 34 | History taking | ANC_PROV_17 | Provider asked about vaginal bleeding in current pregnancy | During the observed ANC visit, the provider took a history of complaints in the current pregnancy and asked about vaginal bleeding. | Not collected in SARA; Collected only in some SPAs (those that include client/provider observation) | BGD 2014 SPA SLE 2017 SARA |  |
| 35 | History taking | ANC_PROV_18 | Provider asked about fever in current pregnancy | During the observed ANC visit, the provider took a history of complaints in the current pregnancy and asked about fever. | Not collected in SARA; Collected only in some SPAs (those that include client/provider observation) | BGD 2014 SPA SLE 2017 SARA |  |
| 36 | History taking | ANC_PROV_19 | Provider asked about headache or blurred vision in current pregnancy | During the observed ANC visit, the provider took a history of complaints in the current pregnancy and asked about headache or blurred vision. | Not collected in SARA; Collected only in some SPAs (those that include client/provider observation) | BGD 2014 SPA SLE 2017 SARA |  |
| 37 | History taking | ANC_PROV_20 | Provider asked about swollen face or hands or extremities in current pregnancy | During the observed ANC visit, the provider took a history of complaints in the current pregnancy and asked about swollen face or hands or extremities. | Not collected in SARA; Collected only in some SPAs (those that include client/provider observation) | BGD 2014 SPA SLE 2017 SARA |  |
| 38 | History taking | ANC_PROV_21 | Provider asked about tiredness or breathlessness in current pregnancy | During the observed ANC visit, the provider took a history of complaints in the current pregnancy and asked about tiredness or breathlessness. | Not collected in SARA; Collected only in some SPAs (those that include client/provider observation) | BGD 2014 SPA SLE 2017 SARA |  |
| 39 | History taking | ANC_PROV_22 | Provider asked about fetal movement (loss of, excessive, normal) in current pregnancy | During the observed ANC visit, the provider took a history of complaints in the current pregnancy and asked about fetal movement (loss of, excessive, normal). | Not collected in SARA; Collected only in some SPAs (those that include client/provider observation) | BGD 2014 SPA SLE 2017 SARA |  |
| 40 | History taking | ANC_PROV_23 | Provider asked about cough or difficulty breathing for 3 weeks or longer in current pregnancy | During the observed ANC visit, the provider took a history of complaints in the current pregnancy and asked about cough or difficulty breathing for 3 weeks or longer. | Not collected in SARA; Collected only in some SPAs (those that include client/provider observation) | BGD 2014 SPA SLE 2017 SARA |  |
| 41 | Examination and observation | ANC_PROV_28 | Observed client for oedema | During the observed ANC visit, the provider examined the client including observing the client for oedema (other than ankle-specific). | Not collected in SARA; Collected only in some SPAs (those that include client/provider observation) | BGD 2014 SPA SLE 2017 SARA |  |
| 42 | Examination and observation | ANC_PROV_30 | Examined client blood pressure | During the observed ANC visit, the provider examined the client including taking a blood pressure measurement. | Not collected in SARA; Collected only in some SPAs (those that include client/provider observation) | BGD 2014 SPA SLE 2017 SARA |  |
| 43 | Examination and observation | ANC_PROV_31 | Examined client weight | During the observed ANC visit, the provider examined the client including taking a weight measurement. | Not collected in SARA; Collected only in some SPAs (those that include client/provider observation) | BGD 2014 SPA SLE 2017 SARA |  |
| 44 | Examination and observation | ANC_PROV_33 | Palpated the clients’ abdomen for fundal height | During the observed ANC visit, the provider examined the client including palpating the clients abdomen for fundal height. | Not collected in SARA; Collected only in some SPAs (those that include client/provider observation) | BGD 2014 SPA SLE 2017 SARA |  |
| 45 | Laboratory investigations | ANC_PROV_37 | Performed or referred the client for hemoglobin testing | During the observed ANC visit, the provider performed or referred the client for hemoglobin testing. | Not collected in SARA; Collected only in some SPAs (those that include client/provider observation) | BGD 2014 SPA SLE 2017 SARA |  |
| 46 | Laboratory investigations | ANC_PROV_38 | Performed or referred the client for grouping and rhesus factor testing | During the observed ANC visit, the provider performed or referred the client for grouping and rhesus factor testing. | Not collected in SARA; Collected only in some SPAs (those that include client/provider observation) | BGD 2014 SPA SLE 2017 SARA |  |
| 47 | Laboratory investigations | ANC_PROV_39 | Performed or referred the client for syphilis testing | During the observed ANC visit, the provider performed or referred the client for syphilis testing. | Not collected in SARA; Collected only in some SPAs (those that include client/provider observation) | BGD 2014 SPA SLE 2017 SARA |  |
| 48 | Laboratory investigations | ANC_PROV_40 | Performed or referred the client for HIV testing | During the observed ANC visit, the provider performed or referred the client for HIV testing. | Not collected in SARA; Collected only in some SPAs (those that include client/provider observation) | BGD 2014 SPA SLE 2017 SARA |  |
| 49 | Laboratory investigations | ANC_PROV_41 | Performed or referred the client for urine- protein, sugar, acetone testing | During the observed ANC visit, the provider performed or referred the client for urine protein, sugar, and acetone testing | Not collected in SARA; Collected only in some SPAs (those that include client/provider observation) | BGD 2014 SPA SLE 2017 SARA |  |
| 50 | Preventative treatment | ANC_PROV_43 | Provided or prescribed iron pills and/or folic acid pills | During the observed ANC visit, the provider prescribed or provided iron pills or folic acid or both | Not collected in SARA; Collected only in some SPAs (those that include client/provider observation) | BGD 2014 SPA SLE 2017 SARA |  |
| 51 | Preventative treatment | ANC_PROV_45 | Provided or prescribed tetanus toxoid immunization | During the observed ANC visit, the provider administered or prescribed tetanus toxoid immunization | Not collected in SARA; Collected only in some SPAs (those that include client/provider observation) | BGD 2014 SPA SLE 2017 SARA |  |
| 52 | Client education and counseling | ANC_PROV_47 | Discussed process of pregnancy and its complications | During the observed ANC visit, the provider discussed the process of pregnancy and its complications | Not collected in SARA; Collected only in some SPAs (those that include client/provider observation) | BGD 2014 SPA SLE 2017 SARA |  |
| 53 | Client education and counseling | ANC_PROV_48 | Discussed diet and nutrition | During the observed ANC visit, the provider discussed diet and nutrition (i.e., quantity and quality of food to eat) during the pregnancy. | Not collected in SARA; Collected only in some SPAs (those that include client/provider observation) | BGD 2014 SPA SLE 2017 SARA |  |
| 54 | Client education and counseling | ANC_PROV_58 | Discussed danger signs in pregnancy | During the observed ANC visit, the provider discussed danger signs in pregnancy. | Not collected in SARA; Collected only in some SPAs (those that include client/provider observation) | BGD 2014 SPA SLE 2017 SARA |  |
| 55 | Client education and counseling | ANC_PROV_49 | Discussed voluntary counselling and testing for HIV | During the observed ANC visit, the provider discussed voluntary counselling and testing for HIV | Not collected in SARA; Collected only in some SPAs (those that include client/provider observation) | BGD 2014 SPA SLE 2017 SARA |  |
| 56 | Client education and counseling | ANC_PROV_50 | Discussed breastfeeding | During the observed ANC visit, the provider discussed early initiation and prolonged breastfeeding as well as exclusive breastfeeding. | Not collected in SARA; Collected only in some SPAs (those that include client/provider observation) | BGD 2014 SPA SLE 2017 SARA |  |
| 57 | Client education and counseling | ANC_PROV_51 | Discussed plans of delivery | During the observed ANC visit, the provider discussed plans of delivery (i.e., emergency preparedness, place of delivery, transportation, financial arrangements). | Not collected in SARA; Collected only in some SPAs (those that include client/provider observation) | BGD 2014 SPA SLE 2017 SARA |  |
| 58 | Client experience | ANC_SATIS_02a | Client satisfied with ability to discuss problems or concerns about pregnancy with provider | Client had no problems with regards to her ability to discuss the problems or concerns about her pregnancy. | Not collected in SARA; Collected only in some SPAs (those that include client/provider observation) | BGD 2014 SPA SLE 2017 SARA |  |
| 59 | Client experience | ANC_SATIS_03a | Client satisfied with the amount of explanation received about the problem or treatment | Client had no problems with regards to the amount of explanation she received about the problem or treatment. | Not collected in SARA; Collected only in some SPAs (those that include client/provider observation) | BGD 2014 SPA SLE 2017 SARA |  |
| 60 | Client experience | ANC_SATIS_10a | Client satisfied with how treated by the staff | Client had no problems with regards to how the staff treated her. | Not collected in SARA; Collected only in some SPAs (those that include client/provider observation) | BGD 2014 SPA SLE 2017 SARA |  |
| 61 | Client experience | ANC_SATIS_04a | Client satisfied with privacy from having others see | Client had no problems with regards to privacy from having others see. | Not collected in SARA; Collected only in some SPAs (those that include client/provider observation) | BGD 2014 SPA SLE 2017 SARA |  |
| 62 | Client experience | ANC_SATIS_05a | Client satisfied with privacy from having others hear | Client had no problems with regards to privacy from having others hear. | Not collected in SARA; Collected only in some SPAs (those that include client/provider observation) | BGD 2014 SPA SLE 2017 SARA |  |
| 63 | Client experience | ANC_SATIS_01a | Client satisfied with time waited | Client had no problems with regards to the time she waited. | Not collected in SARA; Collected only in some SPAs (those that include client/provider observation) | BGD 2014 SPA SLE 2017 SARA |  |
| 64 | Client experience | ANC_SATIS_08a | Client satisfied with number of days services are available | Client had no problems with regards to the number of days services are available. | Not collected in SARA; Collected only in some SPAs (those that include client/provider observation) | BGD 2014 SPA SLE 2017 SARA |  |
| 65 | Client experience | ANC_SATIS_07a | Client satisfied with the hours of service | Client had no problems with regards to the hours of service | Not collected in SARA; Collected only in some SPAs (those that include client/provider observation) | BGD 2014 SPA SLE 2017 SARA |  |
| 66 | Client experience | ANC_SATIS_11a | Client satisfied with the cost for services or treatments | Client had no problems with regards to the cost for services or treatment | Not collected in SARA; Collected only in some SPAs (those that include client/provider observation) | BGD 2014 SPA SLE 2017 SARA |  |
| 67 | Client experience | ANC_SATIS_06a | Client satisfied with the availability of medicines | Client had no problems with regards to the availability of medicines | Not collected in SARA; Collected only in some SPAs (those that include client/provider observation) | BGD 2014 SPA SLE 2017 SARA |  |
| 68 | Client experience | ANC_SATIS_09a | Client satisfied with the cleanliness of the facility | Client had no problems with regards to the cleanliness of the facility | Not collected in SARA; Collected only in some SPAs (those that include client/provider observation) | BGD 2014 SPA SLE 2017 SARA |  |

Supplementary Table 4: Maternal nutrition readiness and provision/experience of care items and detailed definitions

| **No** | **Domain** | **Code** | **Item** | **Detailed indicator definition** | **Data availability notes** | **Survey specific: Indicator not collected** | **Cross-country differences** |
| --- | --- | --- | --- | --- | --- | --- | --- |
| **Readiness** | | | | | | | |
| 1 | Equipment and supplies | ANC_READY_21 | Adult weighing scale | Adult weighing scale observed available and functional in the OPD or ANC service provision area. | SARA only asks about this item in the OPD |  | Sierra Leone is OPD only |
| 2 | Equipment and supplies | ANC_READY_29 | Stadiometer or height rod | Stadiometer or height rod/height board observed available and functional in the OPD or ANC service provision area. | Collected only in some SPAs  SARA only asks about this item in the OPD | BGD 2014 HTI 2013 NPL 2015 SEN 2016 | Sierra Leone is OPD only |
| 3 | Equipment and supplies | GEN_READY_17 | Latex gloves | Clean/sterile disposable latex gloves observed available in the OPD or ANC service provision area. | SARA only asks about this item in the OPD |  | Sierra Leone is OPD only |
| 4 | Equipment and supplies | GEN_READY_16 | Handwashing materials | Either handwashing soap and running water OR alcohol-based hand sanitizer observed available in the OPD or ANC service provision area. | SARA only asks about this item in the OPD |  | Sierra Leone is OPD only |
| 5 | Equipment and supplies | GEN_READY_14 | Environmental disinfectant | Disinfectant (e.g., chlorine, hibitane, alcohol) observed available anywhere in the facility. | SARA only asks about this item in the OPD whereas SPA is in multiple service areas |  | Sierra Leone is OPD only |
| 6 | Equipment and supplies | GEN_READY_15 | Auto-disable syringes with needles; single use standard disposable syringes with needles | Single-use standard disposable syringes with needles or auto-disable syringes with needles observed available anywhere in the facility. | SARA only asks about this item in the OPD whereas SPA is in multiple service areas |  | Sierra Leone is OPD only |
| 7 | Diagnostics | ANC_READY_41 | Blood glucose | Facility has the ability to conduct blood glucose testing onsite and has the required equipment available and functioning/unexpired (glucometer and test strips OR blood chemistry analyzer) |  |  |  |
| 8 | Diagnostics | ANC_READY_04 | Hemoglobin | Facility has the ability to conduct hemoglobin testing onsite and has the required equipment available and functioning/unexpired (rapid test OR hematology analyzer OR hemocue and microcuvette OR colorimeter or hemoglobinometer & drabkin's solution & pipette OR litmus paper for hemoglobin test). |  |  | Haiti, Malawi, Nepal, Senegal, and Bangladesh don't include functioning of microcuvette or drabkin solution unexpired Nepal also doesn't include litmus paper for hemoglobin test Sierra Leone only includes heomcue, colorimeter, hemoglobinometer |
| 9 | Medicines and commodities | ANC_READY_31 | Albendazole/ mebendazole | Albendazole or mebendazole tablets for deworming observed in pharmacy or anywhere in the facility where medicines are routinely stored; at least one with valid expiration date. |  |  |  |
| 10 | Medicines and commodities | ANC_READY_07 | Folic acid tablets | Folic acid tablets or iron + folic acid combination tablets observed in pharmacy or anywhere in the facility where medicines are routinely stored; at least one with valid expiration date. |  |  |  |
| 11 | Medicines and commodities | ANC_READY_06 | Iron tablets | Iron tablets (stand-alone, not in combination with folic acid) observed in pharmacy or anywhere in the facility where medicines are routinely stored; at least one with valid expiration date. |  |  |  |
| 12 | Medicines and commodities | ANC_READY_09 | Sulfadoxine-pyrimethamine (SP) tablets | Sulfadoxine-pyrimethamine (SP) tablets for IPTp observed in pharmacy or anywhere in the facility where medicines are routinely stored; at least one with valid expiration date. | Collected only in some SPAs; malaria endemic countries | BGD 2014 HTI 2013 NPL 2015 |  |
| 13 | Human resources | ANC_READY_01 | Guidelines for antenatal care | Guidelines for antenatal care are observed available in the antenatal care service provision area. |  |  |  |
| 14 | Human resources | ANC_READY_44 | Guidelines for intermittent preventive treatment of malaria during pregnancy | Guidelines for intermittent preventive treatment of malaria during pregnancy are observed available in the antenatal care service provision area. | Collected only in some SPAs; malaria endemic countries | BGD 2014 HTI 2013 NPL 2015 |  |
| 15 | Human resources | ANC_READY_43 | Guidelines for infant and young child feeding | Guidelines for infant and young child feeding are observed available in the PMTCT service provision area. | Collected only in some SPAs; high HIV burden countries | BGD 2014 NPL 2015 |  |
| 16 | Human resources | ANC_READY_02 | Staff trained in the past 2 years in antenatal care (broad ANC training) | Proportion of health care workers delivering ANC services that have been trained in antenatal care (regardless of the specific topic area) in the last two years.  Alternative: At least one health care worker providing ANC services has been trained in antenatal care (regardless of the specific topic area) in the last two years. |  |  | Sierra Leone uses alternative definition while SPA countries use preferred/ original definition |
| 17 | Human resources | ANC_READY_45 | Staff trained in the past 2 years in ANC screening (e.g., blood pressure, urine glucose, and protein) | Proportion of health care workers delivering ANC services that have been trained in ANC screening (e.g., blood pressure, urine glucose, and protein) in the last two years. | Not collected in SARA | SLE 2017 |  |
| 18 | Human resources | ANC_READY_46 | Staff trained in the past 2 years in counseling for ANC (e.g., nutrition, FP, and newborn care) | Proportion of health care workers delivering ANC services that have been trained in ANC screening (e.g., blood pressure, urine glucose, and protein) in the last two years. | Not collected in SARA | SLE 2017 |  |
| 19 | Human resources | ANC_READY_33 | Staff trained in the past 2 years in nutritional assessment of the pregnant woman (e.g., BMI calculation, MUAC measurement) | Proportion of health care workers delivering ANC services that have been trained in nutritional assessment of the pregnant woman (e.g., BMI calculation, MUAC measurement) in the last two years. | Not collected in SARA | SLE 2017 |  |
| 20 | Human resources | ANC_READY_47 | Staff trained in the past 2 years in complications of pregnancy and their management | Proportion of health care workers delivering ANC services that have been trained in complications of pregnancy and their management in the last two years. | Not collected in SARA | SLE 2017 |  |
| 21 | Human resources | ANC_READY_48 | Staff trained in the past 2 years in infant and young child feeding | Proportion of health care workers delivering ANC services that have been trained in infant and young child feeding in the last two years. |  |  | Sierra Leone uses alternative definition while SPA countries use preferred/ original definition |
| 22 | Basic amenities | GEN_READY_40 | Clean environment | General assessment of cleanliness/conditions of the facility; score out of 8 possible good responses including the following eight areas: Floor swept, no obvious dirt, Counters/tables/chairs wiped, Needles/sharps outside of sharps container, Sharps box overflowing/torn/pierced, Bandages/ infectious waste lying uncovered, Walls with significant damage, Doors with significant damage, Ceiling with water stains/ damage | Not collected in SARA | SLE 2017 |  |
| 23 | Basic amenities | GEN_READY_07 | Emergency transportation | Facility has a functioning vehicle with fuel that is routinely available that can be used for emergency transportation or access to a vehicle in near proximity that can be used for emergency transportation |  |  |  |
| 24 | Basic amenities | GEN_READY_02 | Improved water source | Facility has an improved water source that is onsite or within 500meters of the facility. |  |  |  |
| 25 | Basic amenities | ANC_READY_22 | Place for women to sit/lie down | Examination bed observed available in the ANC service provision area. | Not collected in SARA | SLE 2017 |  |
| 26 | Basic amenities | GEN_READY_01 | Power | Facility is connected to the central supply electricity grid and electricity is always available, or facility has a functional backup generator with fuel (or charged battery), or facility has access to solar power |  |  |  |
| 27 | Basic amenities | GEN_READY_04 | Sanitation facilities | Facility has a functioning toilet (latrine) available for general outpatient client use. |  |  |  |
| **Provision/experience of care** | | | | | | | |
| 28 | Laboratory investigations | ANC_PROV_37 | Performed or referred the client for hemoglobin testing | During the observed ANC visit, the provider performed or referred the client for hemoglobin testing. | Not collected in SARA; Collected only in some SPAs (those that include client/provider observation) | BGD 2014 SLE 2017 |  |
| 29 | Examination and observation | ANC_PROV_63 | Inspected conjunctiva or examined the client for pallor | During the observed ANC visit, the provider inspected conjunctiva or examined the client for pallor. | Not collected in SARA; Collected only in some SPAs (those that include client/provider observation) | BGD 2014 SLE 2017 |  |
| 30 | Preventative treatment | ANC_PROV_43 | Provided or prescribed iron pills and/or folic acid pills | During the observed ANC visit, the provider prescribed or provided iron pills or folic acid or both | Not collected in SARA; Collected only in some SPAs (those that include client/provider observation) | BGD 2014 SLE 2017 |  |
| 31 | Client education and counseling | ANC_PROV_65 | Explained the purpose of iron or folic acid | During the observed ANC visit, the provider explained the purpose of iron or folic acid | Not collected in SARA; Collected only in some SPAs (those that include client/provider observation) | BGD 2014 SLE 2017 |  |
| 32 | Client education and counseling | ANC_PROV_66 | Explained how to take iron or folic acid pills | During the observed ANC visit, the provider explained how to take iron or folic acid pills | Not collected in SARA; Collected only in some SPAs (those that include client/provider observation) | BGD 2014 SLE 2017 |  |
| 33 | Client education and counseling | ANC_PROV_68 | Advised on potential side effects of iron or folic acid pills | During the observed ANC visit, the provider advised on potential side effects of iron or folic acid pills | Not collected in SARA; Collected only in some SPAs (those that include client/provider observation) | BGD 2014 SLE 2017 |  |
| 34 | Preventative treatment | ANC_PROV_80 | Provided or prescribed albendazole or mebendazole | During the observed ANC visit, the provider prescribed or provided albendazole or mebendazole | Not collected in SARA; Collected only in some SPAs (those that include client/provider observation) | BGD 2014 SLE 2017 |  |
| 35 | Client education and counseling | ANC_PROV_50a | Discussed exclusive breastfeeding | During the observed ANC visit, the provider discussed exclusive breastfeeding. | Not collected in SARA; Collected only in some SPAs (those that include client/provider observation) | BGD 2014 SLE 2017 |  |
| 36 | Client education and counseling | ANC_PROV_50b | Discussed early initiation and prolonged breastfeeding | During the observed ANC visit, the provider discussed early initiation and prolonged breastfeeding. | Not collected in SARA; Collected only in some SPAs (those that include client/provider observation) | BGD 2014 SLE 2017 |  |
| 37 | Client education and counseling | ANC_PROV_48 | Discussed nutrition during the pregnancy | During the observed ANC visit, the provider discussed nutrition (i.e., quantity and quality of food to eat) during the pregnancy. | Not collected in SARA; Collected only in some SPAs (those that include client/provider observation) | BGD 2014 SLE 2017 |  |
| 38 | Preventative treatment | ANC_PROV_44 | Provided or prescribed sulfadoxine-pyrimethamine | During the observed ANC visit, the provider prescribed or provided sulfadoxine-pyrimethamine | Not collected in SARA; Collected only in some SPAs (those that include client/provider observation; malaria endemic countries) | BGD 2014 SLE 2017 |  |
| 39 | Client experience | ANC_SATIS_03a | Client satisfied with the amount of explanation received about the problem or treatment | Client had no problems with regards to the amount of explanation she received about the problem or treatment. | Not collected in SARA; Collected only in some SPAs (those that include client/provider observation) | BGD 2014 SLE 2017 |  |
| 40 | Client experience | ANC_SATIS_10a | Client satisfied with how treated by the staff | Client had no problems with regards to how the staff treated her. | Not collected in SARA; Collected only in some SPAs (those that include client/provider observation) | BGD 2014 SLE 2017 |  |
| 41 | Client experience | ANC_SATIS_02a | Client satisfied with ability to discuss problems or concerns about pregnancy with provider | Client had no problems with regards to her ability to discuss the problems or concerns about her pregnancy. | Not collected in SARA; Collected only in some SPAs (those that include client/provider observation) | BGD 2014 SLE 2017 |  |

Supplementary Table 5: Overview of indicators and data sources used to define each step of the coverage cascade

| **Step of the coverage cascade** | **Service** | **Indicator(s)** | **Indicator abbreviation** | **Data source(s)** |
| --- | --- | --- | --- | --- |
| Target population | - ANC - Maternal nutrition | Women ages 15-49 with a live birth in the last two years |  | Household survey |
| Service contact | - ANC - Maternal nutrition | Proportion of women ages 15-49 with a live birth in the last two years who had at least 1 ANC contact during the most recent pregnancy | ANC1 | Household survey |
|  |  | Proportion of women ages 15-49 with a live birth in the last two years who had four or more ANC contacts during the most recent pregnancy | ANC4 |  |
|  |  | Proportion of women ages 15-49 with a live birth in the last two years who had 8 or more ANC contacts during the most recent pregnancy | ANC8 |  |
| Readiness-adjusted coverage | - ANC | Proportion of women ages 15-49 with a live birth in the last two years who received ANC from an ANC "ready" facility  Readiness includes:   - ***Basic amenities:*** improved water source, room with auditory and visual privacy, sanitation facilities - ***Diagnostics***: hemoglobin, urine dipstick- protein, urine dipstick- glucose, syphilis RDT / RPR, HIV diagnostic capacity - ***Equipment and supplies***: blood pressure apparatus, stethoscope, examination bed, latex gloves, single use syringes with needles, handwashing materials, environmental disinfectant - ***Human resources***: staff trained in the past 2 years in antenatal care - ***Medicines and commodities***: iron tablets, folic acid tablets, tetanus toxoid vaccine |  | Household survey and Health facility assessment (facility inventory, Health worker interview) |
|  | - Maternal nutrition | Proportion of women ages 15-49 with a live birth in the last two years who received ANC from a nutrition "ready" facility  Readiness includes:   - ***Basic amenities***: clean environment, emergency transportation, improved water source, place for women to sit/lie down, power, sanitation facilities - ***Diagnostics***: blood glucose, hemoglobin - ***Equipment and supplies***: adult weighing scale, stadiometer or height rod, latex gloves, handwashing materials, environmental disinfectant, single use syringes with needles - ***Human resources***: guidelines for antenatal care, guidelines for intermittent preventive treatment of malaria during pregnancy, guidelines for infant and young child feeding, staff trained in the past 2 years in antenatal care, staff trained in the past 2 years in ANC screening, staff trained in the past 2 years in counseling for ANC, staff trained in the past 2 years in nutritional assessment of the pregnant woman, staff trained in the past 2 years in complications of pregnancy and their management, staff trained in the past 2 years in infant and young child feeding - ***Medicines and commodities***: albendazole/ mebendazole, folic acid tablets, iron tablets, sulfadoxine-pyrimethamine |  |  |
| Intervention coverage | - ANC | Proportion of women ages 15-49 with a live birth in the last two years who reported receiving key interventions (average score of interventions received) during the most recent pregnancy  Interventions include: |  | Household survey |
|  |  | - Tetanus toxoid vaccination^ꝉ^   - Received two doses of tetanus toxoid during this pregnancy OR at least 2 doses, the last within 3 years OR at least 3 doses, the last within 5 years OR at least 4 doses, the last within 10 years; OR at least 5 doses during lifetime | TT |  |
|  |  | - SP for IPTp**^§^**   - At least 2 times during pregnancy | SP for IPTp |  |
|  |  | - Iron/folic acid supplementation   - Took at least 90 days of IFA during the pregnancy | IFA |  |
|  |  | - Blood pressure   - Measured at least once during the pregnancy | BP |  |
|  |  | - Urine sample   - Taken at least once during the pregnancy |  |  |
|  |  | - Blood sample   - Taken at least once during the pregnancy |  |  |
|  |  | - Deworming   - Received drugs for intestinal parasites during pregnancy |  |  |
|  | - Maternal nutrition | Proportion of women ages 15-49 with a live birth in the last two years who reported receiving key interventions (average score of interventions received) during the most recent pregnancy  Interventions include: |  |  |
|  |  | - SP for IPTp**^§^**   - At least 2 times during pregnancy | SP for IPTp |  |
|  |  | - Iron/folic acid supplementation   - Took at least 90 days of IFA during the pregnancy | IFA |  |
|  |  | - Blood sample   - Taken at least once during the pregnancy |  |  |
|  |  | - Deworming   - Received drugs for intestinal parasites during pregnancy |  |  |
| Quality-adjusted coverage | - ANC | Proportion of women ages 15-49 with a live birth in the last two years who received ANC services according to standard protocols  Quality includes:  ***History taking:*** Provider asked about client age, Provider asked about medications client is taking, Provider asked about date last menstrual period began, Provider asked about any prior pregnancy, Provider asked about previous still birth, Provider asked about prior pregnancy in which the infant died in the first week of life, Provider asked about heavy bleeding during or after delivery in prior pregnancies, Provider asked about previous assisted delivery, Provider asked about previous spontaneous abortion, Provider asked about previous multiple pregnancies, Provider asked about previous prolonged labor, Provider asked about previous pregnancy-induced hypertension, Provider asked about previous pregnancy related convulsions, Provider asked about high fever or infection during prior pregnancy/ pregnancies, Provider asked about vaginal bleeding in current pregnancy, Provider asked about fever in current pregnancy, Provider asked about headache or blurred vision in current pregnancy, Provider asked about swollen face or hands or extremities in current pregnancy, Provider asked about tiredness or breathlessness in current pregnancy, Provider asked about fetal movement (loss of, excessive, normal) in current pregnancy, Provider asked about cough or difficulty breathing for 3 weeks or longer in current pregnancy  ***Examination and observation:*** Observed client for oedema, Examined client blood pressure, Examined client weight, Palpated the clients' abdomen for fundal height  ***Laboratory investigations:*** Performed or referred the client for hemoglobin testing, Performed or referred the client for grouping and rhesus factor testing, Performed or referred the client for syphilis testing, Performed or referred the client for HIV testing, Performed or referred the client for urine- protein, sugar, acetone testing  ***Preventative treatment:*** Provided or prescribed iron pills and/or folic acid pills, Provided or prescribed tetanus toxoid immunization  ***Client education and counseling:*** Discussed process of pregnancy and its complications, Discussed diet and nutrition, Discussed danger signs in pregnancy, Discussed voluntary counselling and testing for HIV, Discussed breastfeeding, Discussed plans of delivery  ***Client experience:*** Client satisfied with ability to discuss problems or concerns about pregnancy with provider, Client satisfied with the amount of explanation received about the problem or treatment, Client satisfied with how treated by the staff, Client satisfied with privacy from having others see, Client satisfied with privacy from having others hear, Client satisfied with time waited, Client satisfied with number of days services are available, Client satisfied with the hours of service, Client satisfied with the cost for services or treatments, Client satisfied with the availability of medicines, Client satisfied with the cleanliness of the facility |  | Household survey and Health facility assessment (ANC client observation, ANC client exit interview) |
|  | - Maternal nutrition | Proportion of women ages 15-49 with a live birth in the last two years who received maternal nutrition services according to standard protocols  Quality includes:  ***Examination and observation:*** Inspected conjunctiva or examined the client for pallor  ***Laboratory investigations:*** Performed or referred the client for hemoglobin testing  ***Preventative treatment:*** Provided or prescribed albendazole or mebendazole, Provided or prescribed sulfadoxine-pyrimethamine, Provided or prescribed iron pills and/or folic acid pills, Explained the purpose of iron or folic acid, Explained how to take iron or folic acid pills, Advised on potential side effects of iron or folic acid pills  ***Client education and counseling:*** Discussed exclusive breastfeeding, Discussed early initiation and prolonged breastfeeding, Discussed nutrition during the pregnancy  ***Client experience:*** Client satisfied with the amount of explanation received about the problem or treatment, Client satisfied with how treated by the staff, Client satisfied with ability to discuss problems or concerns about pregnancy with provider |  |  |

§ For SP for IPTp, the clinical guidelines were updated in 2013 followed by a change in the global monitoring indicator definition in 2016/2017 prompting us to use the older definition for pre-2016 surveys and the newer definition for post-2016 surveys [70,71]. However, the only post-2016 survey in our analysis did not ask about SP for IPTp thus effectively we only used the older SP for IPTp definition.

ꝉ For TT we used the most recent indicator definitions agreed upon by the international community [72,73].

Supplementary Figure 1: Distribution of ANC readiness scores, by strata and country

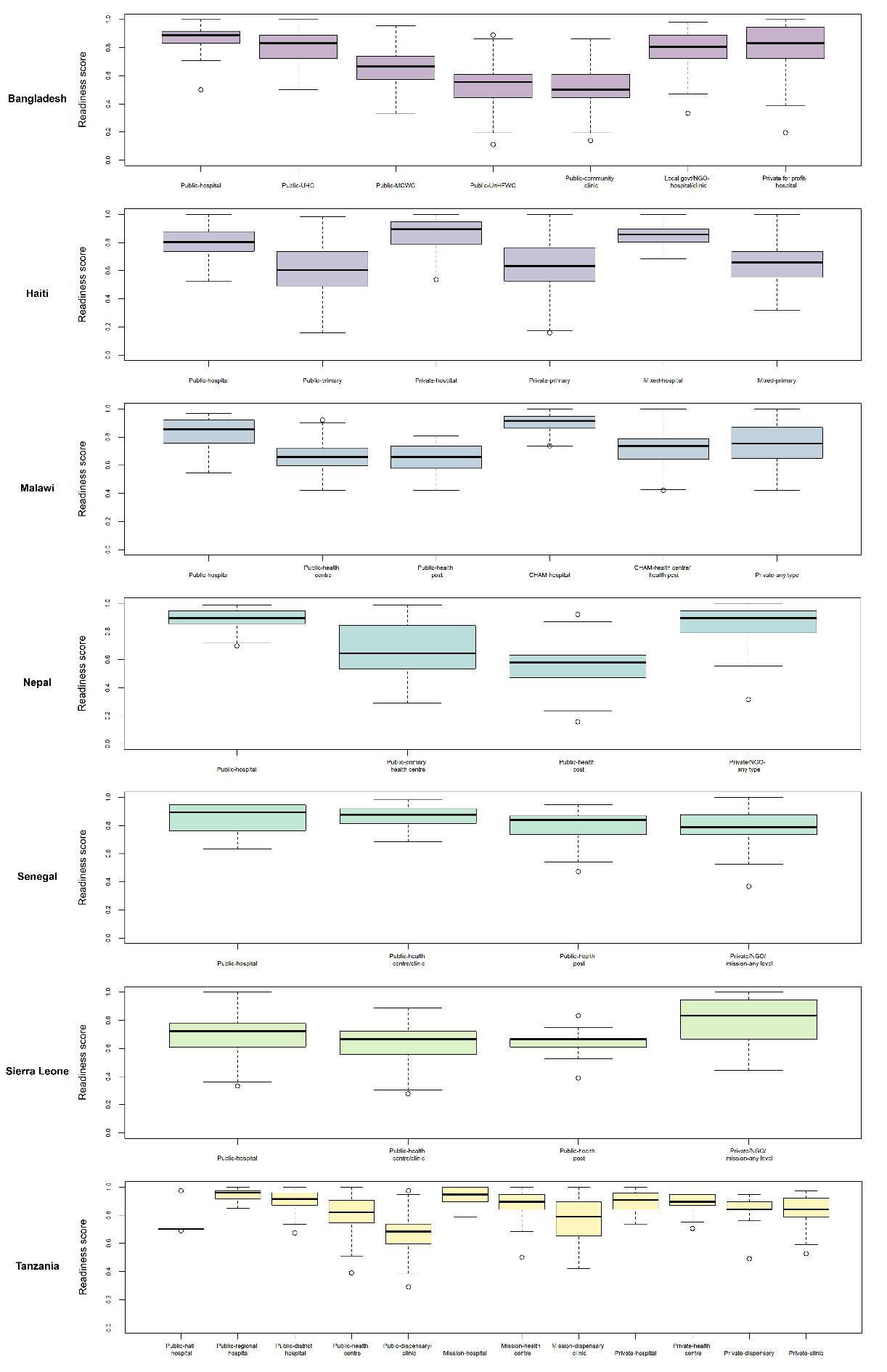

Supplementary Figure 2: Distribution of maternal nutrition readiness scores, by strata and country

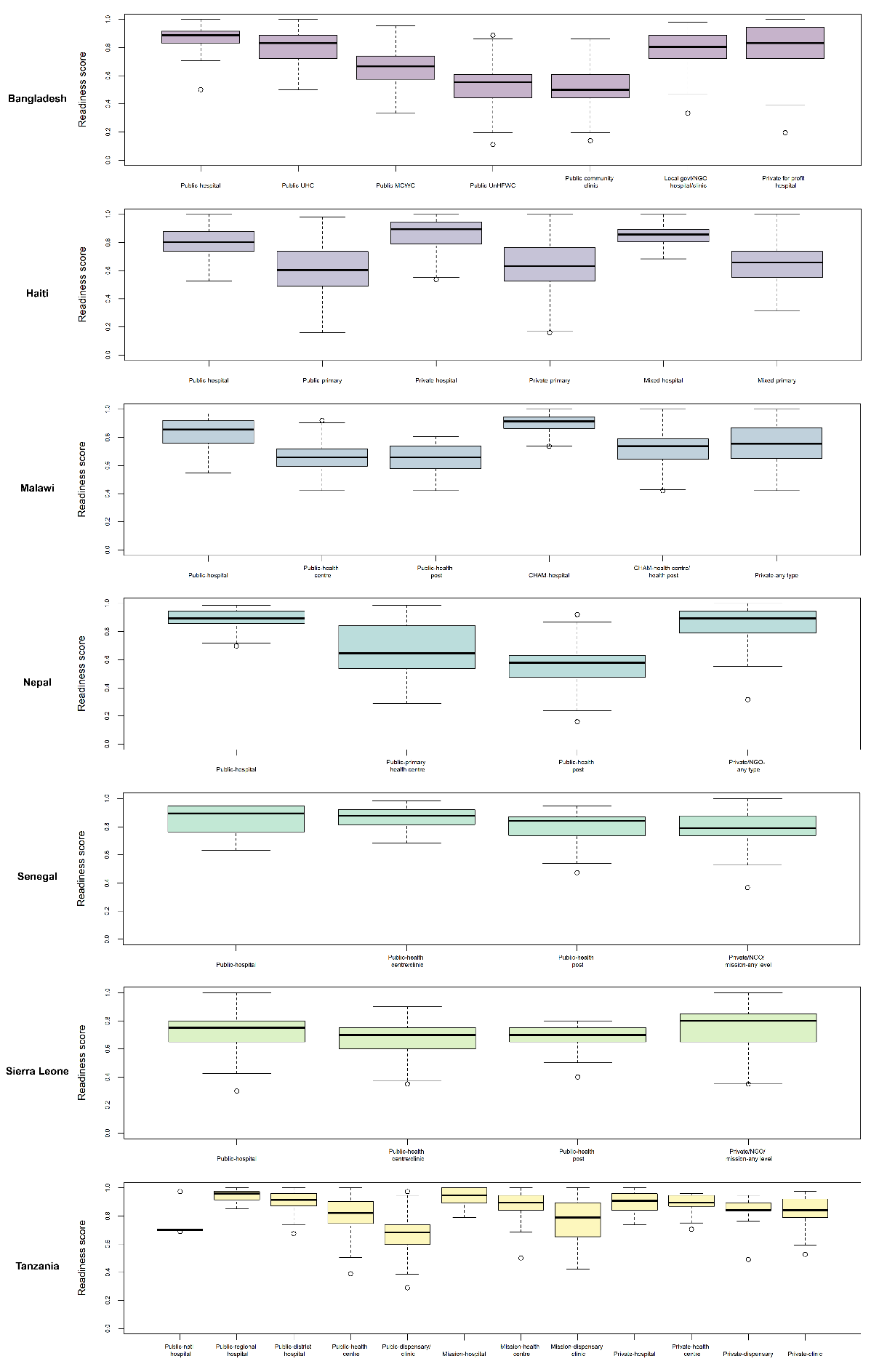

Supplementary Table 6: Availability of ANC readiness items, by country

| **Item** | **Bangladesh**  **N = 1493** | | | **Haiti**  **N = 832** | | | **Malawi**  **N = 643** | | | **Nepal**  **N = 872** | | | **Senegal**  **N = 323** | | | **Sierra Leone**  **N = 1247** | | | **Tanzania**  **N = 1032** | | |
| --- | --- | --- | --- | --- | --- | --- | --- | --- | --- | --- | --- | --- | --- | --- | --- | --- | --- | --- | --- | --- | --- |
|  | **Mean** | **CI_L** | **CI_U** | **Mean** | **CI_L** | **CI_U** | **Mean** | **CI_L** | **CI_U** | **Mean** | **CI_L** | **CI_U** | **Mean** | **CI_L** | **CI_U** | **Mean** | **CI_L** | **CI_U** | **Mean** | **CI_L** | **CI_U** |
| ***Equipment*** | | | | | | | | | | | | | | | | | | | | | |
| Blood pressure apparatus | 87% | 84% | 90% | 95% | 93% | 96% | 68% | 65% | 72% | 86% | 83% | 89% | 98% | 96% | 100% | 80% | 78% | 83% | 79% | 75% | 83% |
| Stethoscope | 92% | 90% | 94% | 98% | 97% | 99% | 75% | 71% | 78% | 89% | 86% | 92% | 94% | 91% | 97% | 92% | 91% | 94% | 83% | 80% | 87% |
| Examination bed | 78% | 74% | 81% | 91% | 89% | 93% | 97% | 96% | 99% | 92% | 90% | 95% | 100% | 100% | 100% |  |  |  | 98% | 97% | 100% |
| Latex gloves | 64% | 60% | 68% | 84% | 82% | 87% | 96% | 94% | 97% | 89% | 86% | 92% | 99% | 98% | 100% | 99% | 98% | 99% | 90% | 88% | 93% |
| Single use syringes | 82% | 78% | 85% | 90% | 88% | 92% | 98% | 97% | 99% | 96% | 94% | 97% | 80% | 75% | 85% | 99% | 98% | 99% | 95% | 93% | 97% |
| Soap and water OR alcohol-based hand rub | 56% | 52% | 60% | 73% | 70% | 76% | 71% | 68% | 75% | 58% | 53% | 63% | 98% | 96% | 100% | 91% | 90% | 93% | 73% | 70% | 77% |
| Disinfectant | 59% | 55% | 63% | 82% | 80% | 85% | 90% | 88% | 93% | 80% | 76% | 84% | 93% | 90% | 96% | 87% | 86% | 89% | 87% | 84% | 90% |
| ***Diagnostics*** | | | | | | | | | | | | | | | | | | | | | |
| Hemoglobin | 12% | 10% | 13% | 33% | 30% | 36% | 25% | 22% | 29% | 14% | 12% | 16% | 12% | 10% | 14% | 4% | 3% | 5% | 30% | 26% | 33% |
| Urine dipstick- protein | 18% | 16% | 21% | 39% | 35% | 42% | 20% | 17% | 23% | 16% | 13% | 18% | 89% | 85% | 92% | 22% | 20% | 24% | 33% | 29% | 37% |
| Urine dipstick- glucose | 18% | 15% | 21% | 39% | 35% | 42% | 18% | 15% | 21% | 14% | 12% | 15% | 86% | 81% | 90% | 18% | 16% | 20% | 29% | 25% | 32% |
| Syphilis RDT / RPR | 6% | 5% | 6% | 37% | 34% | 41% | 29% | 25% | 32% | 11% | 10% | 13% | 4% | 2% | 6% | 5% | 3% | 6% | 52% | 48% | 57% |
| HIV testing / RDT |  |  |  | 37% | 33% | 40% | 95% | 93% | 96% | 15% | 13% | 17% | 91% | 88% | 95% | 21% | 16% | 27% | 90% | 88% | 93% |
| ***Medicines*** | | | | | | | | | | | | | | | | | | | | | |
| Iron tablets | 96% | 94% | 97% | 82% | 80% | 85% | 97% | 96% | 98% | 92% | 90% | 94% | 83% | 79% | 88% | 61% | 59% | 95% | 94% | 93% | 96% |
| Folic acid tablets | 94% | 92% | 96% | 75% | 72% | 78% | 94% | 92% | 96% | 92% | 89% | 94% | 82% | 77% | 86% | 89% | 87% | 79% | 95% | 94% | 97% |
| TT vaccine | 19% | 16% | 22% | 35% | 32% | 39% | 77% | 74% | 80% | 25% | 22% | 29% | 92% | 89% | 95% | 88% | 86% | 83% | 86% | 83% | 89% |
| ***Basic amenities and Human resources*** | | | | | | | | | | | | | | | | | | | | | |
| Improved water source | 87% | 84% | 90% | 69% | 66% | 73% | 95% | 93% | 96% | 82% | 78% | 85% | 95% | 93% | 98% | 82% | 80% | 70% | 65% | 61% | 69% |
| Room with auditory and visual privacy | 29% | 26% | 33% | 74% | 71% | 77% | 78% | 75% | 82% | 67% | 63% | 72% | 59% | 54% | 65% | 57% | 54% | 96% | 73% | 69% | 76% |
| Sanitation facilities | 87% | 84% | 90% | 81% | 78% | 84% | 90% | 88% | 93% | 90% | 87% | 93% | 100% | 99% | 100% | 71% | 68% | 84% | 66% | 62% | 70% |
| Proportion of staff offering ANC trained in ANC in last two years | 16% | 14% | 19% | 35% | 33% | 37% | 37% | 35% | 40% | 10% | 9% | 12% | 68% | 62% | 73% | 83% | 81% | 97% | 34% | 32% | 37% |
| ***Overall score*** | | | | | | | | | | | | | | | | | | | | | |
| Readiness score | 55% | 54% | 56% | 66% | 65% | 67% | 71% | 70% | 72% | 59% | 58% | 60% | 80% | 79% | 81% | 87% | 85% | 89% | 71% | 70% | 72% |

Supplementary Table 7: Availability of maternal nutrition readiness items, by country

| **Item** | **Bangladesh**  **N = 1493** | | | **Haiti**  **N = 832** | | | **Malawi**  **N = 643** | | | **Nepal**  **N = 889** | | | **Senegal**  **N = 323** | | | **Sierra Leone**  **N = 1247** | | | **Tanzania**  **N = 1031** | | |
| --- | --- | --- | --- | --- | --- | --- | --- | --- | --- | --- | --- | --- | --- | --- | --- | --- | --- | --- | --- | --- | --- |
|  | **Mean** | **CI_L** | **CI_U** | **Mean** | **CI_L** | **CI_U** | **Mean** | **CI_L** | **CI_U** | **Mean** | **CI_L** | **CI_U** | **Mean** | **CI_L** | **CI_U** | **Mean** | **CI_L** | **CI_U** | **Mean** | **CI_L** | **CI_U** |
| ***Equipment*** |  |  |  |  |  |  |  |  |  |  |  |  |  |  |  |  |  |  |  |  |  |
| Adult weighing scale | 83% | 80% | 86% | 86% | 83% | 88% | 88% | 86% | 91% | 87% | 83% | 90% | 97% | 95% | 99% | 61% | 58% | 64% | 84% | 81% | 87% |
| Stadiometer/height rod |  |  |  |  |  |  | 60% | 56% | 64% |  |  |  |  |  |  |  |  |  | 61% | 57% | 65% |
| Environmental disinfectant | 59% | 55% | 63% | 82% | 80% | 85% | 90% | 88% | 93% | 80% | 76% | 83% | 93% | 90% | 96% | 87% | 86% | 89% | 87% | 84% | 90% |
| Single-use syringes with needles | 82% | 78% | 85% | 90% | 88% | 92% | 98% | 97% | 99% | 96% | 94% | 97% | 80% | 75% | 85% | 99% | 98% | 99% | 95% | 93% | 97% |
| Handwashing materials | 56% | 52% | 60% | 73% | 70% | 76% | 71% | 68% | 75% | 58% | 53% | 62% | 98% | 96% | 100% | 91% | 90% | 93% | 73% | 70% | 77% |
| Latex gloves | 64% | 60% | 68% | 84% | 82% | 87% | 96% | 94% | 97% | 89% | 86% | 92% | 99% | 98% | 100% | 99% | 98% | 99% | 90% | 88% | 93% |
| ***Diagnostics*** |  |  |  |  |  |  |  |  |  |  |  |  |  |  |  |  |  |  |  |  |  |
| Hemoglobin | 12% | 10% | 13% | 33% | 30% | 36% | 25% | 22% | 29% | 14% | 12% | 15% | 12% | 10% | 14% | 4% | 3% | 5% | 30% | 26% | 33% |
| Blood glucose | 8% | 6% | 9% | 37% | 34% | 41% | 21% | 18% | 25% | 7% | 6% | 8% | 84% | 80% | 88% | 5% | 4% | 7% | 18% | 15% | 20% |
| ***Medicines*** |  |  |  |  |  |  |  |  |  |  |  |  |  |  |  |  |  |  |  |  |  |
| Iron tablets | 62% | 58% | 66% | 65% | 62% | 68% | 37% | 34% | 41% | 12% | 10% | 15% | 6% | 3% | 8% | 84% | 82% | 86% | 41% | 37% | 45% |
| Folic acid tablets | 94% | 92% | 96% | 75% | 72% | 78% | 94% | 92% | 96% | 92% | 89% | 94% | 82% | 77% | 86% | 79% | 77% | 81% | 95% | 94% | 97% |
| IPT drug (SP) |  |  |  |  |  |  | 99% | 98% | 100% |  |  |  | 75% | 70% | 80% | 82% | 80% | 84% | 61% | 57% | 65% |
| De-worming drugs (Mebendazole/Albendazole) | 88% | 85% | 91% | 88% | 86% | 90% | 98% | 96% | 99% | 97% | 95% | 98% | 0% | 0% | 1% | 88% | 86% | 90% | 90% | 88% | 93% |
| ***Basic amenities*** |  |  |  |  |  |  |  |  |  |  |  |  |  |  |  |  |  |  |  |  |  |
| Place for women to sit/lie down (examination bed) | 78% | 74% | 81% | 91% | 89% | 93% | 97% | 96% | 99% | 92% | 90% | 95% | 100% | 100% | 100% |  |  |  | 98% | 97% | 100% |
| Power | 21% | 18% | 24% | 65% | 61% | 68% | 67% | 63% | 70% | 44% | 40% | 48% | 58% | 52% | 64% | 52% | 49% | 55% | 64% | 60% | 69% |
| Improved water source | 87% | 84% | 90% | 69% | 66% | 73% | 95% | 93% | 96% | 81% | 77% | 85% | 95% | 93% | 98% | 57% | 54% | 59% | 65% | 61% | 69% |
| Sanitation facilities | 87% | 84% | 90% | 81% | 78% | 84% | 90% | 88% | 93% | 90% | 86% | 93% | 100% | 99% | 100% | 83% | 81% | 85% | 66% | 62% | 70% |
| Emergency transportation | 9% | 7% | 10% | 37% | 34% | 40% | 83% | 80% | 86% | 57% | 52% | 61% | 64% | 59% | 70% | 91% | 90% | 93% | 59% | 55% | 64% |
| Clean environment | 72% | 70% | 74% | 91% | 90% | 92% | 89% | 88% | 90% | 82% | 79% | 84% | 93% | 92% | 95% |  |  |  | 82% | 80% | 84% |
| ***Human resources*** |  |  |  |  |  |  |  |  |  |  |  |  |  |  |  |  |  |  |  |  |  |
| Guidelines ANC | 50% | 46% | 54% | 33% | 30% | 37% | 59% | 55% | 63% | 25% | 21% | 29% | 77% | 72% | 82% | 62% | 59% | 65% | 56% | 51% | 60% |
| Proportion of staff offering ANC trained in ANC in last two years | 16% | 14% | 19% | 35% | 33% | 37% | 37% | 35% | 40% | 10% | 9% | 12% | 68% | 62% | 73% | 87% | 85% | 89% | 34% | 32% | 37% |
| Proportion of staff offering ANC trained in nutritional assessment of pregnant women in last two years | 10% | 8% | 13% | 13% | 12% | 15% | 11% | 9% | 13% | 5% | 3% | 7% | 29% | 25% | 34% |  |  |  | 6% | 4% | 7% |
| Guidelines IYCF |  |  |  | 10% | 8% | 12% | 30% | 27% | 34% |  |  |  | 26% | 21% | 31% | 38% | 36% | 41% | 33% | 29% | 37% |
| Guidelines IPTP |  |  |  |  |  |  | 49% | 45% | 53% |  |  |  | 67% | 61% | 72% | 64% | 62% | 67% | 35% | 31% | 39% |
| Proportion of staff offering ANC trained in ANC screening in last two years | 14% | 11% | 16% | 14% | 12% | 16% | 11% | 9% | 13% | 8% | 6% | 10% | 37% | 32% | 42% |  |  |  | 8% | 6% | 10% |
| Proportion of staff offering ANC trained in counseling for ANC in last two years | 16% | 13% | 19% | 16% | 14% | 18% | 13% | 11% | 15% | 8% | 6% | 10% | 38% | 32% | 43% |  |  |  | 8% | 6% | 10% |
| Proportion of staff offering ANC trained in complications of pregnancy and their management in last two years | 14% | 12% | 17% | 15% | 13% | 17% | 14% | 11% | 16% | 8% | 6% | 11% | 32% | 27% | 37% |  |  |  | 8% | 6% | 10% |
| Proportion of staff offering ANC trained in infant and young child feeding in last two years | 10% | 7% | 12% | 10% | 9% | 12% | 26% | 23% | 28% | 3% | 2% | 4% | 15% | 12% | 19% |  |  |  | 27% | 24% | 30% |
| ***Overall score*** |  |  |  |  |  |  |  |  |  |  |  |  |  |  |  |  |  |  |  |  |  |
| Readiness score | 47% | 46% | 48% | 54% | 53% | 55% | 61% | 60% | 62% | 50% | 49% | 51% | 63% | 61% | 64% | 70% | 70% | 71% | 55% | 54% | 56% |

Supplementary Figure 3: Distribution of ANC provision/experience of care scores, by strata and country

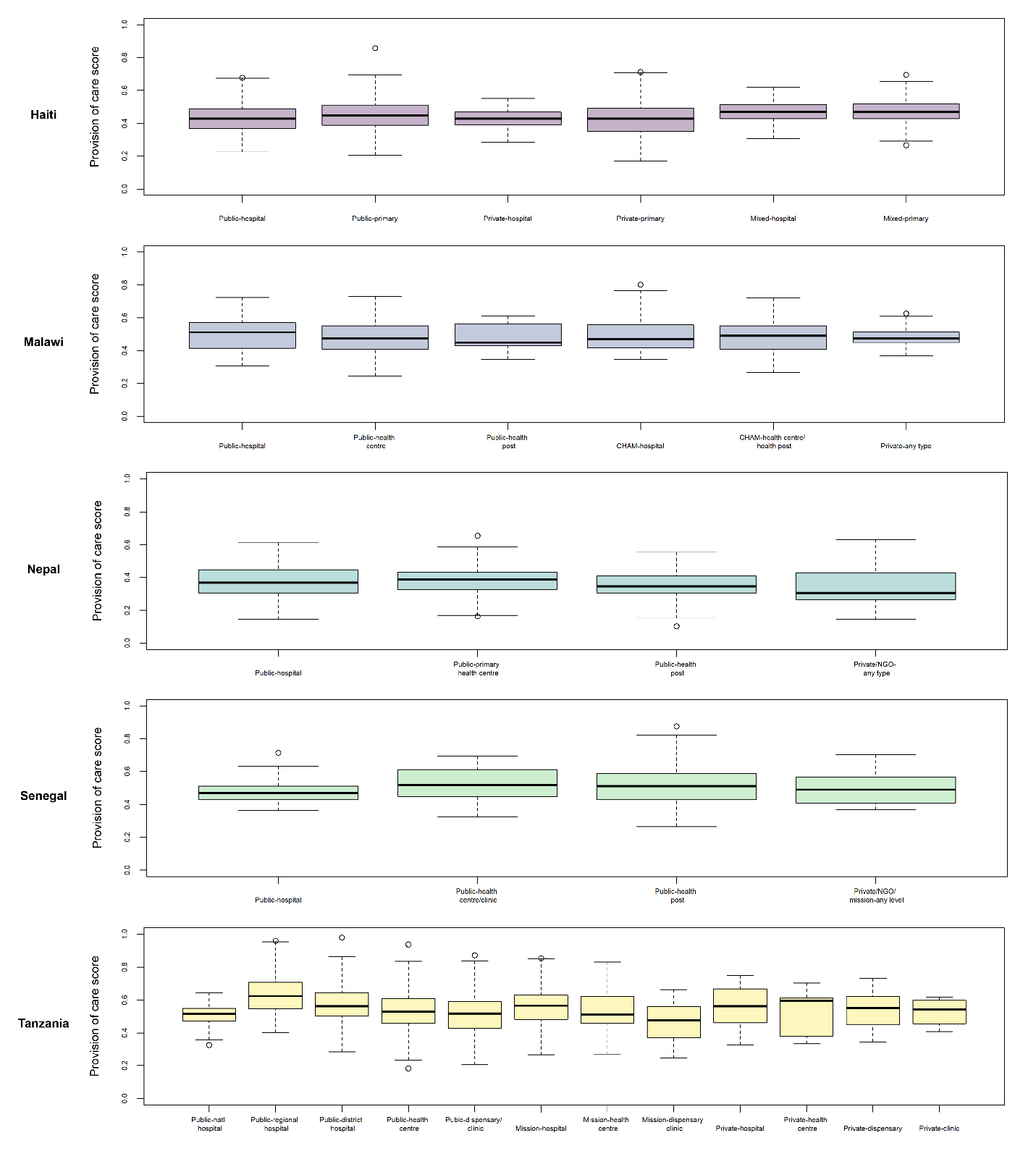

Supplementary Figure 4: Distribution of maternal nutrition provision/experience of care scores, by strata and country

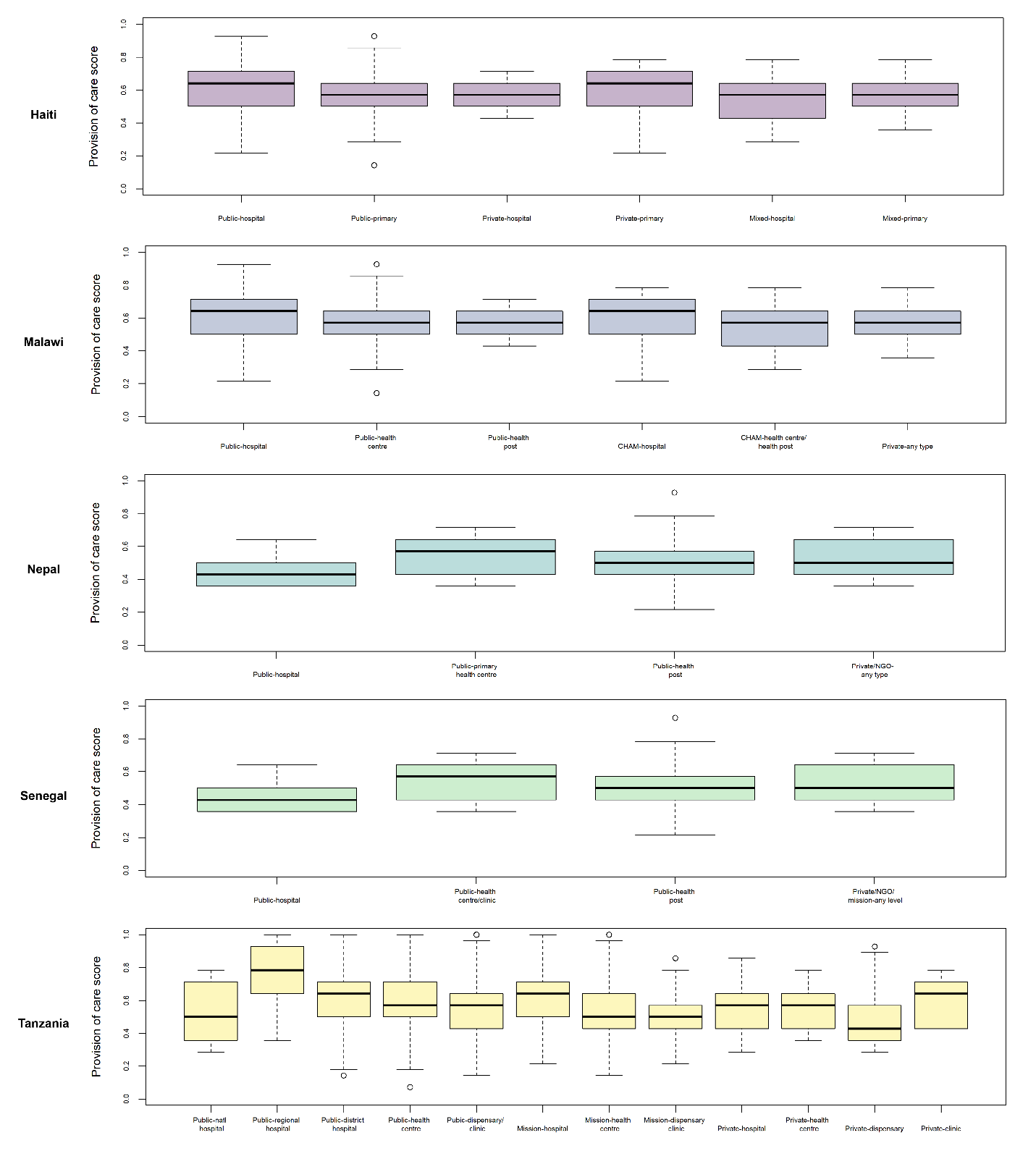

Supplementary Table 8: Availability of ANC provision/experience of care items, by country

| **Item** | **Haiti**  **N = 785** | | | **Malawi**  **N = 859** | | | **Nepal**  **N = 573** | | | **Senegal**  **N = 307** | | | **Tanzania**  **N = 1737** | | |
| --- | --- | --- | --- | --- | --- | --- | --- | --- | --- | --- | --- | --- | --- | --- | --- |
|  | **Mean** | **CI_L** | **CI_U** | **Mean** | **CI_L** | **CI_U** | **Mean** | **CI_L** | **CI_U** | **Mean** | **CI_L** | **CI_U** | **Mean** | **CI_L** | **CI_U** |
| ***History taking: Personal and Past medical*** | | | | | | | | | | | | | | | |
| Personal history: client age | 72% | 69% | 76% | 80% | 77% | 83% | 54% | 48% | 59% | 91% | 87% | 95% | 91% | 89% | 93% |
| Personal history: medications client is taking | 11% | 9% | 13% | 12% | 9% | 15% | 12% | 8% | 15% | 12% | 8% | 17% | 12% | 10% | 14% |
| Personal history: date last menstrual period began | 84% | 81% | 86% | 76% | 73% | 79% | 78% | 72% | 83% | 73% | 67% | 79% | 81% | 78% | 84% |
| Personal history: any prior pregnancy | 73% | 69% | 76% | 89% | 86% | 91% | 63% | 58% | 69% | 89% | 85% | 93% | 87% | 84% | 89% |
| Past medical history for prior pregnancies: still birth | 25% | 22% | 29% | 43% | 39% | 47% | 6% | 4% | 8% | 48% | 41% | 54% | 28% | 25% | 31% |
| Past medical history for prior pregnancies: infant died in the first week of life | 9% | 6% | 11% | 23% | 20% | 26% | 1% | 0% | 2% | 14% | 9% | 19% | 24% | 21% | 26% |
| Past medical history for prior pregnancies: heavy bleeding during or after delivery | 4% | 2% | 6% | 37% | 33% | 41% | 1% | 0% | 1% | 4% | 2% | 7% | 25% | 22% | 28% |
| Past medical history for prior pregnancies: previous assisted delivery | 12% | 9% | 14% | 47% | 43% | 51% | 3% | 2% | 4% | 6% | 3% | 9% | 29% | 26% | 32% |
| Past medical history for prior pregnancies: previous spontaneous abortion | 24% | 20% | 27% | 46% | 42% | 50% | 8% | 5% | 10% | 20% | 14% | 25% | 37% | 34% | 41% |
| Past medical history for prior pregnancies: previous multiple pregnancies | 9% | 7% | 12% | 22% | 19% | 25% | 1% | 0% | 2% | 63% | 56% | 69% | 6% | 4% | 7% |
| Past medical history for prior pregnancies: previous prolonged labor | 1% | 0% | 1% | 8% | 6% | 10% | 1% | 0% | 3% | 21% | 16% | 27% | 4% | 3% | 6% |
| Past medical history for prior pregnancies: pregnancy-induced hypertension | 8% | 6% | 10% | 29% | 26% | 33% | 1% | 0% | 1% | 3% | 0% | 5% | 14% | 12% | 17% |
| Past medical history for prior pregnancies: pregnancy related convulsions | 3% | 1% | 4% | 26% | 23% | 30% | 1% | 0% | 1% | 15% | 10% | 19% | 3% | 2% | 5% |
| Past medical history for prior pregnancies: high fever or infection during prior pregnancy | 2% | 1% | 4% | 3% | 2% | 4% | 0% | 0% | 0% | 4% | 2% | 7% | 2% | 1% | 3% |
| ***History taking: Complaints in pregnancy*** | | | | | | | | | | | | | | | |
| History of complaints in current pregnancy: vaginal bleeding | 14% | 11% | 16% | 26% | 23% | 30% | 9% | 6% | 11% | 3% | 1% | 4% | 50% | 47% | 53% |
| History of complaints in current pregnancy: fever | 13% | 11% | 16% | 11% | 8% | 13% | 7% | 4% | 11% | 3% | 1% | 5% | 25% | 22% | 27% |
| History of complaints in current pregnancy: headache or blurred vision | 31% | 28% | 35% | 18% | 16% | 21% | 14% | 10% | 18% | 16% | 11% | 21% | 42% | 39% | 45% |
| History of complaints in current pregnancy: swollen face or hands or extremities | 5% | 4% | 7% | 12% | 10% | 14% | 19% | 14% | 24% | 1% | 0% | 2% | 31% | 28% | 34% |
| History of complaints in current pregnancy: tiredness or breathlessness | 9% | 7% | 11% | 5% | 4% | 7% | 3% | 2% | 5% | 6% | 3% | 9% | 20% | 18% | 23% |
| History of complaints in current pregnancy: fetal movement (loss of, excessive, normal) | 8% | 6% | 10% | 22% | 19% | 25% | 19% | 15% | 22% | 11% | 8% | 15% | 32% | 29% | 35% |
| History of complaints in current pregnancy: cough or difficulty breathing for 3 weeks or longer | 7% | 5% | 9% | 7% | 5% | 9% | 1% | 0% | 1% | 1% | 0% | 3% | 5% | 4% | 7% |
| ***Observation and clinical investigation*** | | | | | | | | | | | | | | | |
| Edema | 20% | 17% | 23% | 65% | 61% | 68% | 26% | 21% | 31% | 86% | 81% | 91% | 59% | 55% | 62% |
| Blood pressure | 93% | 91% | 95% | 48% | 44% | 52% | 83% | 79% | 88% | 99% | 97% | 100% | 72% | 69% | 75% |
| Weight | 81% | 78% | 84% | 74% | 70% | 77% | 72% | 67% | 78% | 97% | 94% | 99% | 79% | 76% | 82% |
| Palpate the client’s abdomen for fundal height | 57% | 54% | 61% | 92% | 91% | 94% | 17% | 13% | 20% | 67% | 61% | 73% | 79% | 76% | 81% |
| ***Laboratory investigation and Drug administration*** | | | | | | | | | | | | | | | |
| Hemoglobin | 59% | 56% | 63% | 10% | 7% | 12% | 35% | 29% | 40% | 40% | 34% | 47% | 47% | 44% | 49% |
| Grouping and rhesus factor | 37% | 33% | 41% | 2% | 1% | 3% | 32% | 27% | 37% | 36% | 30% | 43% | 34% | 31% | 36% |
| RPR (syphilis test) | 53% | 49% | 56% | 13% | 10% | 15% | 19% | 15% | 22% | 44% | 37% | 50% | 42% | 39% | 45% |
| HIV testing | 53% | 49% | 57% | 53% | 49% | 56% | 16% | 12% | 20% | 65% | 58% | 71% | 71% | 68% | 74% |
| Urine- protein, sugar, acetone | 55% | 52% | 59% | 4% | 2% | 6% | 32% | 27% | 37% | 65% | 59% | 71% | 41% | 39% | 44% |
| Iron and/or folic acid provided or prescribed | 73% | 70% | 77% | 90% | 88% | 92% | 71% | 66% | 77% | 95% | 92% | 98% | 83% | 81% | 86% |
| Tetanus toxoid provided or prescribed | 25% | 22% | 28% | 45% | 41% | 49% | 31% | 27% | 36% | 65% | 59% | 72% | 52% | 49% | 55% |
| ***Client education and counselling*** | | | | | | | | | | | | | | | |
| Process of pregnancy and its complications | 18% | 15% | 21% | 57% | 53% | 60% | 24% | 19% | 30% | 23% | 18% | 29% | 50% | 47% | 53% |
| Diet and nutrition | 45% | 41% | 49% | 44% | 41% | 48% | 53% | 47% | 59% | 43% | 36% | 49% | 35% | 31% | 38% |
| Danger signs in pregnancy | 6% | 5% | 7% | 14% | 12% | 16% | 2% | 1% | 3% | 18% | 14% | 21% | 36% | 34% | 37% |
| Voluntary counselling and testing for HIV | 15% | 12% | 18% | 41% | 37% | 45% | 7% | 5% | 10% | 64% | 58% | 71% | 75% | 72% | 78% |
| Breastfeeding | 7% | 5% | 9% | 10% | 7% | 13% | 2% | 1% | 4% | 2% | 1% | 4% | 16% | 13% | 18% |
| Plans of delivery (emergency preparedness, place of delivery, transportation, financial arrangements) | 34% | 30% | 37% | 82% | 79% | 85% | 13% | 9% | 17% | 20% | 15% | 26% | 69% | 66% | 72% |
| ***Experience of care*** | | | | | | | | | | | | | | | |
| No problem: discuss problems or concerns | 90% | 88% | 92% | 93% | 91% | 95% | 80% | 75% | 84% | 99% | 99% | 100% | 96% | 95% | 97% |
| No problem: explanation you received | 92% | 90% | 94% | 96% | 94% | 97% | 78% | 73% | 82% | 100% | 100% | 100% | 96% | 95% | 97% |
| No problem: how the staff treated you | 97% | 96% | 98% | 94% | 92% | 96% | 96% | 94% | 98% | 100% | 99% | 100% | 96% | 95% | 97% |
| No problem: privacy from having others see | 93% | 91% | 95% | 99% | 98% | 99% | 84% | 80% | 89% | 100% | 100% | 100% | 96% | 95% | 97% |
| No problem: privacy from having others hear | 92% | 90% | 94% | 98% | 97% | 99% | 85% | 81% | 89% | 100% | 100% | 100% | 96% | 95% | 97% |
| No problem: time you waited | 72% | 68% | 75% | 73% | 70% | 76% | 66% | 60% | 71% | 77% | 72% | 83% | 69% | 65% | 72% |
| No problem: number of days services are available | 93% | 91% | 95% | 91% | 89% | 93% | 90% | 86% | 94% | 98% | 97% | 100% | 92% | 90% | 94% |
| No problem: hours of service | 94% | 93% | 96% | 85% | 83% | 88% | 91% | 88% | 94% | 97% | 96% | 99% | 90% | 89% | 92% |
| No problem: cost for services or treatments | 93% | 91% | 95% | 92% | 90% | 93% | 92% | 89% | 95% | 97% | 95% | 99% | 94% | 92% | 95% |
| No problem: availability of medicines | 90% | 88% | 92% | 86% | 83% | 88% | 87% | 82% | 91% | 93% | 90% | 96% | 78% | 75% | 80% |
| No problem: cleanliness of the facility | 93% | 92% | 95% | 92% | 91% | 94% | 87% | 84% | 91% | 98% | 96% | 100% | 85% | 83% | 88% |
| ***Overall score*** | | | | | | | | | | | | | | | |
| Provision/experience of care score | 44% | 43% | 45% | 49% | 48% | 49% | 36% | 35% | 37% | 51% | 50% | 52% | 53% | 52% | 53% |

Supplementary Table 9: Availability of nutrition provision/experience of care items, by country

| **Item** | **Haiti**  **N = 785** | | | **Malawi**  **N = 859** | | | **Nepal**  **N = 573** | | | **Senegal**  **N = 307** | | | **Tanzania**  **N = 1754** | | |
| --- | --- | --- | --- | --- | --- | --- | --- | --- | --- | --- | --- | --- | --- | --- | --- |
|  | **Mean** | **CI_L** | **CI_U** | **Mean** | **CI_L** | **CI_U** | **Mean** | **CI_L** | **CI_U** | **Mean** | **CI_L** | **CI_U** | **Mean** | **CI_L** | **CI_U** |
| ***Observation, laboratory investigation, and drug administration*** | | | | | | | | | | | | | | | |
| Hemoglobin | 59% | 56% | 63% | 10% | 7% | 12% | 35% | 29% | 40% | 40% | 34% | 47% | 47% | 44% | 49% |
| Inspected conjunctiva or examined the client for pallor | 52% | 48% | 56% | 82% | 79% | 85% | 34% | 29% | 40% | 96% | 94% | 99% | 62% | 59% | 65% |
| Antimalarials (SP) provided or prescribed |  |  |  | 75% | 71% | 78% |  |  |  | 34% | 27% | 40% | 47% | 44% | 51% |
| Albendazole or mebendazole provided or prescribed | 0% | 0% | 1% | 71% | 67% | 74% | 47% | 41% | 53% | 18% | 13% | 23% | 55% | 52% | 58% |
| Iron and/or folic acid provided or prescribed | 73% | 70% | 77% | 90% | 88% | 92% | 71% | 66% | 77% | 95% | 92% | 98% | 83% | 81% | 86% |
| Provider gave counseling on purpose of iron tablets | 28% | 25% | 32% | 65% | 61% | 68% | 26% | 21% | 32% | 46% | 40% | 53% | 65% | 62% | 68% |
| Provider gave counseling on how to take iron tablets | 35% | 31% | 38% | 64% | 61% | 68% | 31% | 25% | 36% | 49% | 42% | 55% | 67% | 64% | 70% |
| Provider gave counseling on side effects of iron tablets | 3% | 1% | 4% | 10% | 8% | 12% | 5% | 3% | 7% | 3% | 1% | 4% | 12% | 10% | 14% |
| ***Client education and counselling*** | | | | | | | | | | | | | | | |
| Diet and nutrition | 45% | 41% | 49% | 44% | 41% | 48% | 53% | 47% | 59% | 43% | 36% | 49% | 35% | 31% | 38% |
| Exclusive breastfeeding | 7% | 5% | 9% | 10% | 7% | 12% | 1% | 0% | 2% | 2% | 0% | 3% | 15% | 12% | 17% |
| Early initiation and prolonged breastfeeding | 7% | 5% | 9% | 10% | 7% | 12% | 1% | 0% | 2% | 1% | 0% | 3% | 15% | 12% | 17% |
| ***Experience of care*** | | | | | | | | | | | | | | | |
| No problem: discuss problems or concerns | 90% | 88% | 92% | 93% | 91% | 95% | 80% | 75% | 84% | 99% | 99% | 100% | 96% | 95% | 97% |
| No problem: explanation you received | 92% | 90% | 94% | 96% | 94% | 97% | 78% | 73% | 82% | 100% | 100% | 100% | 96% | 95% | 97% |
| No problem: how the staff treated you | 97% | 96% | 98% | 94% | 92% | 96% | 96% | 94% | 98% | 100% | 99% | 100% | 96% | 95% | 97% |
| ***Overall score*** | | | | | | | | | | | | | | | |
| Provision/experience of care score | 45% | 44% | 46% | 58% | 57% | 59% | 43% | 41% | 45% | 52% | 50% | 54% | 57% | 55% | 58% |

Supplementary Figure 5: ANC8 effective coverage cascades, by country

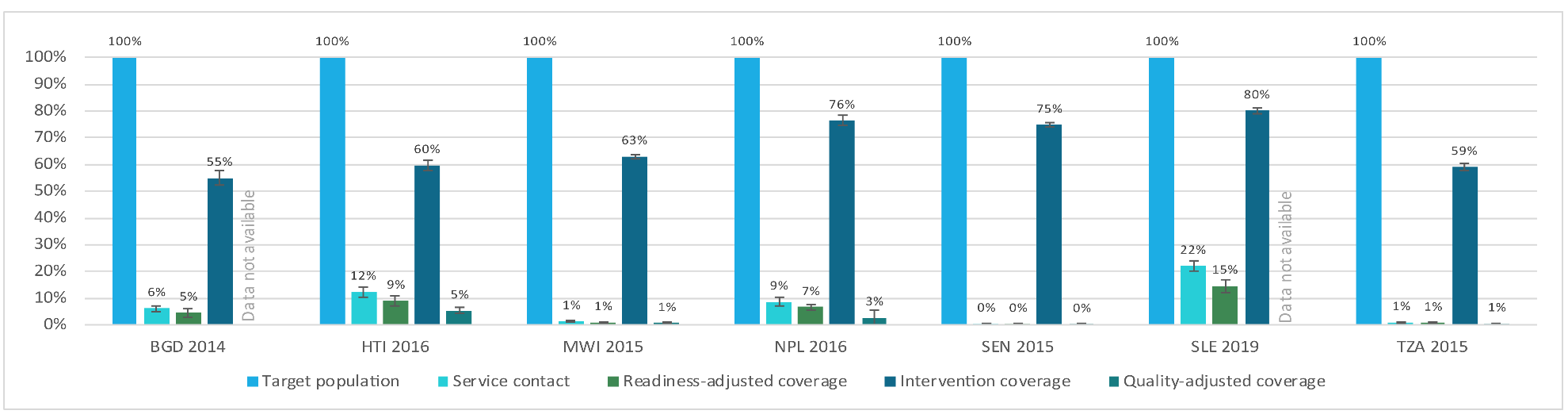

Supplementary Table 10: ANC effective coverage cascade estimates, by country

|  | **Target population** | **Service contact** | | **Readiness-adjusted coverage** | | **Intervention coverage** | | **Quality-adjusted coverage** | |
| --- | --- | --- | --- | --- | --- | --- | --- | --- | --- |
|  | Women with a live birth in the last two years | Received at least 1/4/8 ANC visit(s) | | Received ANC1/4/8 from a "ready" facility | | Received ANC component interventions | | Received ANC1/4/8 services according to standard protocols | |
| **ANC1** |  | **%** | **CI** | **%** | **CI** | **%** | **CI** | **%** | **CI** |
| BGD 2014 | 100% | 80% | (77% - 83%) | 59% | (55% - 63%) | 55% | (52% - 58%) |  |  |
| HTI 2016 | 100% | 90% | (89% - 92%) | 64% | (61% - 67%) | 60% | (58% - 62%) | 39% | (37% - 41%) |
| MWI 2015 | 100% | 98% | (98% - 99%) | 69% | (68% - 71%) | 63% | (62% - 64%) | 47% | (46% - 49%) |
| NPL 2016 | 100% | 96% | (95% - 97%) | 68% | (63% - 73%) | 76% | (75% - 78%) | 33% | (30% - 36%) |
| SEN 2015 | 100% | 98% | (98% - 99%) | 78% | (76% - 79%) | 75% | (74% - 76%) | 48% | (47% - 50%) |
| SLE 2019 | 100% | 99% | (98% - 99%) | 66% | (64% - 67%) | 80% | (79% - 81%) |  |  |
| TZA 2015 | 100% | 98% | (97% - 98%) | 72% | (70% - 74%) | 59% | (58% - 60%) | 51% | (50% - 53%) |
| **ANC4** |  |  |  |  |  |  |  |  |  |
| BGD 2014 | 100% | 31% | (29% - 34%) | 23% | (19% - 26%) | 55% | (52% - 58%) |  |  |
| HTI 2016 | 100% | 63% | (60% - 66%) | 45% | (41% - 49%) | 60% | (58% - 62%) | 27% | (25% - 30%) |
| MWI 2015 | 100% | 48% | (46% - 50%) | 34% | (33% - 36%) | 63% | (62% - 64%) | 23% | (22% - 24%) |
| NPL 2016 | 100% | 71% | (68% - 74%) | 51% | (47% - 54%) | 76% | (75% - 78%) | 24% | (19% - 29%) |
| SEN 2015 | 100% | 56% | (53% - 58%) | 44% | (42% - 47%) | 75% | (74% - 76%) | 27% | (26% - 29%) |
| SLE 2019 | 100% | 79% | (78% - 81%) | 53% | (50% - 56%) | 80% | (79% - 81%) |  |  |
| TZA 2015 | 100% | 48% | (46% - 50%) | 36% | (33% - 39%) | 59% | (58% - 60%) | 26% | (23% - 28%) |
| **ANC8** |  |  |  |  |  |  |  |  |  |
| BGD 2014 | 100% | 6% | (5% - 7%) | 5% | (3% - 6%) | 55% | (52% - 58%) |  |  |
| HTI 2016 | 100% | 12% | (11% - 14%) | 9% | (7% - 11%) | 60% | (58% - 62%) | 5% | (4% - 6%) |
| MWI 2015 | 100% | 1% | (1% - 2%) | 1% | (1% - 1%) | 63% | (62% - 64%) | 1% | (0% - 1%) |
| NPL 2016 | 100% | 9% | (7% - 10%) | 7% | (6% - 8%) | 76% | (75% - 78%) | 3% | (0% - 5%) |
| SEN 2015 | 100% | 0% | (0% - 1%) | 0% | (0% - 1%) | 75% | (74% - 76%) | 0% | (0% - 0%) |
| SLE 2019 | 100% | 22% | (20% - 24%) | 15% | (12% - 17%) | 80% | (79% - 81%) |  |  |
| TZA 2015 | 100% | 1% | (1% - 1%) | 1% | (0% - 1%) | 59% | (58% - 60%) | 1% | (0% - 1%) |

Supplementary Figure 6: ANC8 maternal nutrition effective coverage cascades, by country

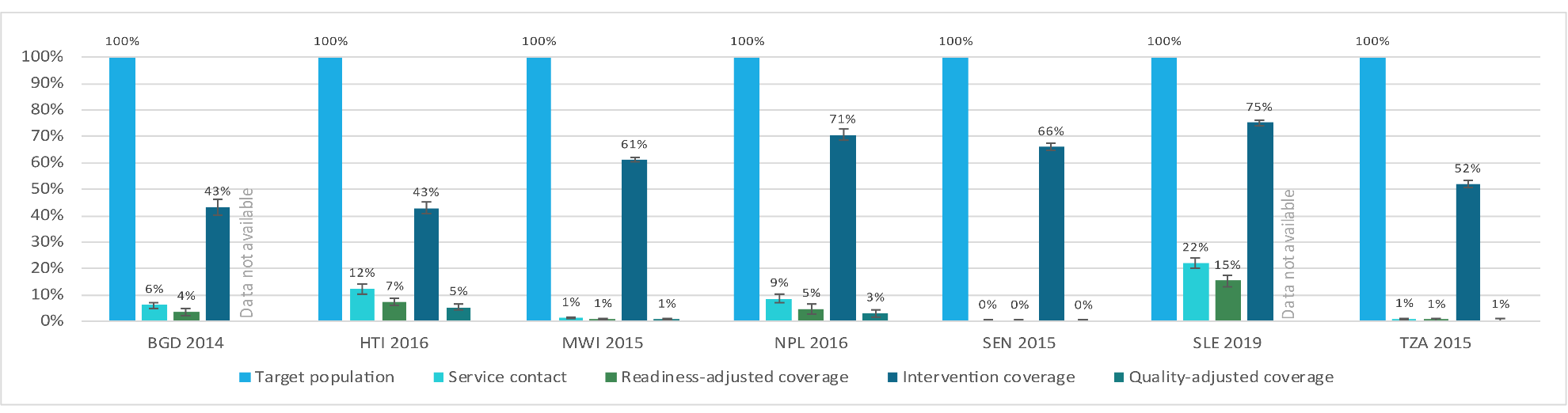

Supplementary Table 11: Maternal nutrition effective coverage cascade estimates, by country

|  | **Target population** | **Service contact** | | **Readiness-adjusted coverage** | | **Intervention coverage** | | **Quality-adjusted coverage** | |
| --- | --- | --- | --- | --- | --- | --- | --- | --- | --- |
|  | Women with a live birth in the last two years | Received at least 1/4/8 ANC visit(s) | | Received ANC1/4/8 from a nutrition "ready" facility | | Received nutrition component interventions | | Received ANC1/4/8 services according to standard protocols for maternal nutrition | |
| **ANC1** |  | **%** | **CI** | **%** | **CI** | **%** | **CI** | **%** | **CI** |
| BGD 2014 | 100% | 80% | (77% - 83%) | 47% | (43% - 50%) | 43% | (40% - 46%) |  |  |
| HTI 2016 | 100% | 90% | (89% - 92%) | 52% | (50% - 55%) | 43% | (41% - 45%) | 40% | (38% - 42%) |
| MWI 2015 | 100% | 98% | (98% - 99%) | 60% | (59% - 62%) | 61% | (60% - 62%) | 56% | (54% - 58%) |
| NPL 2016 | 100% | 96% | (95% - 97%) | 51% | (49% - 54%) | 71% | (68% - 73%) | 40% | (34% - 45%) |
| SEN 2015 | 100% | 98% | (98% - 99%) | 62% | (60% - 64%) | 66% | (65% - 67%) | 50% | (46% - 53%) |
| SLE 2019 | 100% | 99% | (98% - 99%) | 69% | (67% - 70%) | 75% | (74% - 76%) |  |  |
| TZA 2015 | 100% | 98% | (97% - 98%) | 55% | (53% - 56%) | 52% | (51% - 53%) | 56% | (54% - 58%) |
| **ANC4** |  |  |  |  |  |  |  |  |  |
| BGD 2014 | 100% | 31% | (29% - 34%) | 18% | (15% - 21%) | 43% | (40% - 46%) |  |  |
| HTI 2016 | 100% | 63% | (60% - 66%) | 37% | (34% - 40%) | 43% | (41% - 45%) | 28% | (26% - 30%) |
| MWI 2015 | 100% | 48% | (46% - 50%) | 30% | (28% - 31%) | 61% | (60% - 62%) | 27% | (26% - 29%) |
| NPL 2016 | 100% | 71% | (68% - 74%) | 38% | (35% - 42%) | 71% | (68% - 73%) | 29% | (24% - 34%) |
| SEN 2015 | 100% | 56% | (53% - 58%) | 35% | (33% - 37%) | 66% | (65% - 67%) | 28% | (26% - 31%) |
| SLE 2019 | 100% | 79% | (78% - 81%) | 56% | (53% - 58%) | 75% | (74% - 76%) |  |  |
| TZA 2015 | 100% | 48% | (46% - 50%) | 28% | (25% - 30%) | 52% | (51% - 53%) | 28% | (25% - 30%) |
| **ANC8** |  |  |  |  |  |  |  |  |  |
| BGD 2014 | 100% | 6% | (5% - 7%) | 4% | (2% - 5%) | 43% | (40% - 46%) |  |  |
| HTI 2016 | 100% | 12% | (11% - 14%) | 7% | (6% - 9%) | 43% | (41% - 45%) | 5% | (4% - 7%) |
| MWI 2015 | 100% | 1% | (1% - 2%) | 1% | (1% - 1%) | 61% | (60% - 62%) | 1% | (0% - 1%) |
| NPL 2016 | 100% | 9% | (7% - 10%) | 5% | (3% - 7%) | 71% | (68% - 73%) | 3% | (2% - 4%) |
| SEN 2015 | 100% | 0% | (0% - 1%) | 0% | (0% - 0%) | 66% | (65% - 67%) | 0% | (0% - 0%) |
| SLE 2019 | 100% | 22% | (20% - 24%) | 15% | (13% - 18%) | 75% | (74% - 76%) |  |  |
| TZA 2015 | 100% | 1% | (1% - 1%) | 1% | (0% - 1%) | 52% | (51% - 53%) | 1% | (0% - 1%) |

Supplementary Figure 7: Comparing ANC and maternal nutrition readiness-adjusted and quality-adjusted coverage, by country

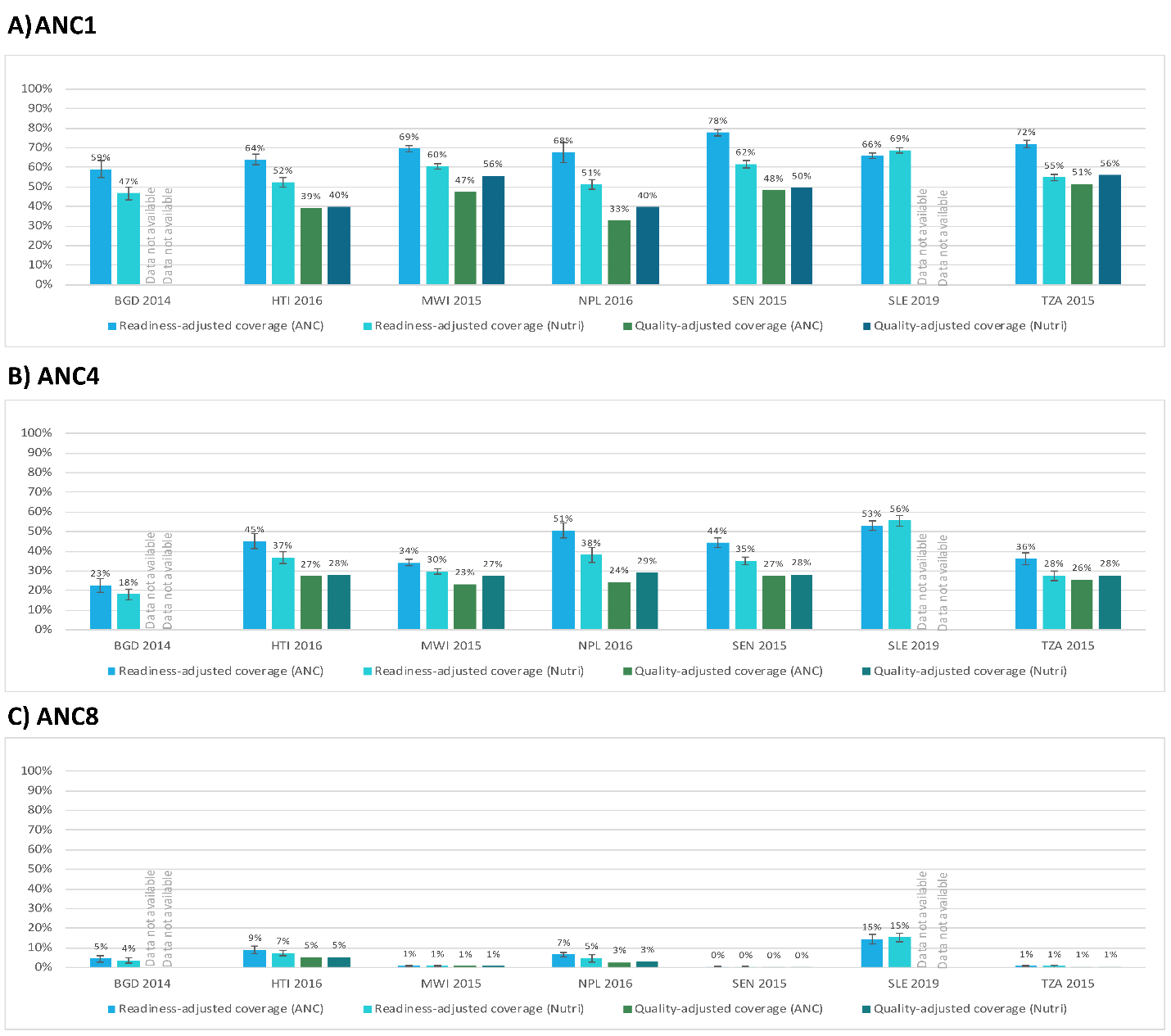
